# SGLT2 inhibitors improved clinical outcomes on biliary diseases beyond glycemic control in patients with type 2 diabetes

**DOI:** 10.1101/2024.10.15.24315461

**Authors:** Ming Gao, Qiuyu Lin, Tingyi Zhu, Kaiyue Hu, Bei Zhong, Kaini Zhang, Xiaoli Chen, Xinyu Chen, Ying Zhang, Yangyang Li, Shaowen Tang, Dongming Su, Xiubin Liang, Yu Liu

## Abstract

**Background & aims:** Diabetes is known to increase the risk of gallstone disease. This study assesses the impact of sodium-glucose cotransporter-2 inhibitors (SGLT2i) on the incidence of biliary diseases, relative to sulfonylureas, in patients with type 2 diabetes mellitus (T2DM) and lithogenic diet (LD)-fed mice.

**Methods:** A retrospective cohort analysis was performed on T2DM patient data who commenced SGLT2i or sulfonylurea therapy from January 1, 2017, to September 1, 2022-sourced from Nanjing Medical University’s database. They were matched using propensity scores (PS) and inverse probability of treatment weighting (IPTW). Follow-up for developing biliary diseases was conducted up to the earliest relevant end-point. Cox models, PS matching, and sensitivity analyses, including standard mortality ratio weighting (SMRW), were applied to determine hazard ratios (HRs) and confidence intervals (CIs). Parallelly, LD-fed C57BL/6J mice were administered SGLT2i or sulfonylureas to corroborate findings in animal models.

**Results:** From the 1,901 patients analyzed over an average of 2.83 years, SGLT2i therapy correlated with a substantially lower risk of developing biliary diseases (HR 0.595, 95% CI 0.410-0.863), particularly among defined subgroups. A downward trend in risk was observed with extended use beyond two years. Concordant data from the mouse model pointed towards SGLT2i mitigating gallstone formation, with putative mechanisms including reduced liver injury and dyslipidemia, as well as improved gallbladder motility and bile acid production.

**Conclusion:** SGLT2i potentially reduces the risk of biliary diseases compared to sulfonylureas, meriting further clinical investigation.

## Introduction

Biliary diseases (BD) include conditions such as gallstones (cholelithiasis), inflammation of the gallbladder (cholecystitis), bile duct inflammation (cholangitis), biliary obstruction due to strictures or tumors, primary sclerosing cholangitis (PSC), primary biliary cholangitis (PBC), biliary cirrhosis, and biliary dyskinesia, which affects the motility of the bile ducts and gallbladder, are prevalent, leading to significant rates of hospitalizations and healthcare expenditures.^1^ A recent meta-analysis, involving 115 studies and 32,610,568 participants, estimated the global prevalence of gallstones at 6.1% (95% confidence intervals (CI), 5.6-6.5) and the incidence at 0.47 per 100 person-years, both increasing with age.^2^ Diabetes is a recognized risk factor for gallstone, where individuals with diabetes exhibit a higher prevalence (15.4%) compared to non-diabetics,^3^ coupled with stronger associations noted in those with higher fasting serum insulin levels and more severe insulin intolerance.

Despite various treatments of glycemic control in type 2 diabetes mellitus (T2DM), the prevalence of BD remains high in T2DM patients. Reducing the incidence of BD is critical in this population. Compared to non-diabetic individuals, T2DM patients are at a substantially higher risk of developing BD,^4^ due to metabolic changes such as insulin resistance, dyslipidemia, and obesity, which contribute to altered bile composition and gallstone formation. Selecting antidiabetic drugs that lower the risk of BD can prevent complications, reduce the need for surgical interventions, and improve patients’ quality of life. Notably, data obtained from recent population-based cohort studies and pairwise and network meta-analyses have indicated that certain treatments, including insulin, dipeptidyl peptidase 4 inhibitors (DPP4i), and glucagon like peptide-1 receptor agonists (GLP-1RA), may even increase the risk of bile duct and gallbladder diseases, including cholelithiasis, in patients with T2DM.^5–7^ These observations point to the need for interventions that target pathways beyond glucose homeostasis to prevent gallstone development.

Sodium-glucose cotransporter-2 inhibitors (SGLT2i) offer a novel approach to diabetes management by promoting glucose excretion in urine. In addition to their impact on glucose maintenance, SGLT2i may confer benefits through several putative mechanisms, including antioxidant, anti-inflammatory, and anti-fibrotic effects.^8^ Recently, an increasing number of clinical trials in patients with or without T2DM indicate that SGLT2i not only improves glycemic control but also exhibits cardiovascular and renal protection^9–11^. Ongoing and future investigations will undoubtedly expand the spectrum of patients who may benefit from SGLT2i.

Recently, a research finding has shown that SGLT2i could reduce the risk of kidney stones in patients with T2DM.^12^ This prompts us to investigate the possible association between the use of SGLT2i and cholelithiasis. We, therefore, investigated SGLT2i’s association with biliary diseases in T2DM patients through a population-based cohort study. Moreover, to investigate its potential mechanisms beyond glycaemic control, we explored the effects of SGLT2i in a lithogenic diet (LD)-fed mice model to consider its prophylactic use in non-diabetic individuals at risk for biliary diseases.

## Materials and Methods

### Human study design

We initially conducted a population-based cohort study including a total of 2.36 million T2DM patients from 2017 to 2022. Patients who met the following criteria were included: (1) patients with T2DM; (2) patients with newly treated with SGLT2i (including canagliflozin, empagliflozin, and dapagliflozin) or sulfonylurea (including glipizide, gliclazide, gliquidone, glimepiride, and glyburide) from January 1, 2017 to September 1, 2022; (3) patients with complete hospitalization information. Patients were excluded if they met any of the following criteria: (1) patients with age under eighteen years or over seventy-five years; (2) pre-menopausal women; (3) patients with estimated glomerular filtration rate (eGFR) under forty-five mL/min/1.73m^2^; (4) patients who have ever used DPP4i or GLP-1RA (which have already been reported to be associated with the occurrence of biliary diseases); (5) patients with prevalent biliary diseases cases; (6) patients with an uncertain diagnosis of biliary diseases during the study period; (7) patients with incomplete information during the study; (8) patients who were lack of medical information before the index date; (9) patients who was treated with both SGLT2i and sulfonylureas; (10) patients who took less than one course of targeted medication; (11) patients who using other types of hypoglycemic drugs in the index date. All these patients were treated in four hospitals of Nanjing Medical University (NMU) in Nanjing, China (Sir Run Run Hospital of NMU, the Affiliated Eye Hospital of NMU, the Geriatric Hospital affiliated to NMU, and the Fourth Affiliated Hospital of NMU). The indnew users did not have a prescription for either SGLT2i or sulfonylureas within six months before the index date was the date of first prescription of either SGLT2i or sulfonylureas, and new users did not have a prescription for either SGLT2i or sulfonylureas within six months before the index date.The outcome is defined as the first occurrence of biliary diseases, including gallstones (cholelithiasis), inflammation of the gallbladder (cholecystitis), bile duct inflammation (cholangitis), biliary obstruction due to strictures or tumors, PSC, PBC, biliary cirrhosis, and biliary dyskinesia, as diagnosed by International Classification of Diseases, Ninth or Tenth Revision, Clinical Modification (ICD-nine/ten-CM) codes (Table S1). Endpoints were: diagnosis of biliary diseases; switch from SGLT2i to sulfonylureas or vice versa; SGLT2i enhancement with sulfonylureas or vice versa; last clinical visit; death record, and end of the entire study. The adapted Diabetes Complications and Severity Index (aDCSI) was used to evaluate the complications and severity of the diabetic patients,^13^ while the age-adjusted Charlson Comorbidity Index (aCCI) score shown in Table S2, was included as a composite comorbidity score to describe patient’s disease burdens,^14^ Medications are demonstrated in Table S3.

### Animal studies

After a one-week adaptive feeding, seven-week-old male C57BL/6J mice were divided into four groups, including a normal diet (ND)-fed group and the other three groups fed with a lithogenic diet (LD) containing 40% high fat, 1.25% cholesterol, and 0.5% cholic acid. These groups also received different treatments (0.76 mg/kg henagliflozin, 0.6 mg/kg glimepiride, and placebo) via oral gavage for eight weeks. Weekly records were maintained for food and water consumption, fasting blood glucose (FBG), and weight. Gallstone formation in mice was monitored every two weeks using Ultrasonic diagnostic instruments and an intraperitoneal glucose (2 mg/kg) tolerance test (IPGTT) was conducted monthly. Prior to sacrifice, all mice underwent an 8-hour fast, and samples of blood, liver, gallbladder, ileum content, and colon contents were collected for analysis.

### Biochemical analysis of serum and bile

The serum biochemistry was examined using an automatic biochemical analyzer, which included measurements for alanine transaminase (ALT), aspartate aminotransferase (AST), alkaline phosphatase (ALP), gamma-glutamyl transpeptidase (GGT), total bile acid (TBA), total bilirubin (TBIL), direct bilirubin (DBIL), indirect bilirubin (IBIL), total cholesterol (TC), triglyceride (TG), low density lipoprotein cholesterol (LDLC), and high density lipoprotein cholesterol (HDLC). Bile samples biochemically for TBA, TC, and LDLC.

### Gallbladder contraction and gallstone analysis

We investigated the potential protective effect of SGLT2i on gallbladder motility by assessing gallbladder contraction in response to intravenous injection of sulfated Cholecystokinin (CCK) Octapeptide in mice following 8 weeks of treatment. The fasting volume of the mice was assessed by ultrasound equipment before injection and again 20 minutes after injection. The gallbladder emptying function was expressed by the following formula: (fasting volume-postprandial volume)/fasting volume*100%.^15^ Gallstone grading was performed according to Akiyoshi’s criteria.^16^

### Liver histopathology

Paraffin-embedded liver sections (5 μm in thickness) were used for hematoxylin and eosin staining (HE staining) and frozen slices (5 μm in thickness) were used for Oil Red O staining to examine liver pathology.

### Cell culture and treatment

Primary hepatocytes were isolated from eight-week-old male C57BL/6J mice using liver perfusion according to our previously published protocol,^17^ and human hepatocytes were obtained from ATCC. Hepatocytes were plated in six-well plates in Dulbecco’s modified Eagle’s medium (DMEM) medium containing 10% fetal bovine serum (FBS) in a 5% CO_2_ humidified incubator at 37°C. In some experiments, cells were stimulated with oleic acid (0.5 mM) for 24 h and treated with 10 μM T-1095 (an inhibitor of SGLT2) for 24 h. Then, the cells were collected for subsequent analysis.

### RNA extraction and quantitative qRT-PCR analysis

RNA was extracted from liver, ilium or cells with RNA-easy Isolation Reagent according to the manufacturer’s instructions. Reverse transcription was performed with Cham Q SYBR qPCR Master Mix and analyzed using a Roche Real-Time PCR System. The relative expression levels of mRNAs were calculated with 2^−ΔΔCt^ method and normalized to the housekeeping gene β-Actin. The primers are listed in Table S4 and other regents and resources are shown in Table S5.

### Statistical analyses

For human studies, to mitigate the impact of confounding factors, we employed propensity score (PS) with inverse probability of treatment weighting (IPTW) to create more comparable groups.^18–20^ Absolute standardized mean differences (ASMD) depicted in Figure S1 were computed to assess differences between comparison groups, with ASMD>0.1 indicating notable distinctions.^21^ We determined the incidence density of biliary diseases per 1000 person-years. Cox model, PS matching (PSM, caliper value 0.05), and sensitivity analyses, including standard mortality ratio weighting (SMRW),^18, 20^ to verify the robustness of results. Additionally, subgroup analyses (including sex, age, presence of diabetic complications, clinical comorbidity status and types of SGLT2i) were conducted to furnish more precise estimates of biliary diseases risk with SGLT2i. All analyses were performed using R software.

For mice study and cell experiments, the statistical significance of differences between the means of experimental groups was evaluated using one-way analysis of variance with a post hoc Bonferroni multiple-comparison test using Prism 9.00 (GraphPad, La Jolla, CA, USA). A difference was considered to be statistically significant at p < 0.05.

### Ethics

This cohort study was approved by Ethic Committee of Sir Run Run Hospital of Nanjing Medical University (2024-SR-040) and the animal study was approved by Institutional Animal Care and Use Committee (IACUC) of Nanjing medical university (IACUC-2308036).

All authors had access to the study data and reviewed and approved the final manuscript.

## Results

### Characteristics of T2DM patients treated with SGLT2i or sulfonylureas

Our database contained records for 37,184 patients with T2DM from January 1, 2017, to September 1, 2022. Out of these, 1,901 individuals met the criteria for inclusion in our study, with 453 initiating SGLT2i and 1,448 initiating sulfonylureas (SU) as shown in Figure 1. Both groups were free of biliary tract diseases at baseline. The mean follow-up period was 2.83 years. We compiled baseline characteristics of the participants, which are presented as counts and percentages, or as means with standard deviations (SD), in Table S6. The age of new users of SGLT2i had a mean (SD) of 59.64 (10.3) years and the adjusted Charlson Comorbidity Index (aCCI) score average (SD) was 5.67 (2.0). These statistics were comparable to those of new sulfonylurea users, who had a mean (SD) age of 62.04 (9.2) years and a mean (SD) aCCI score of 5.55 (1.8). In the SGLT2i group, 61.9% of patients had at least one diabetic complication, a percentage that was closely paralleled by the 55.4% in the sulfonylureas group. After adjusting for the inverse probability of treatment weighting (IPTW)-based propensity scores and excluding the top and bottom 1% of extreme values, there was no significant difference in baseline characteristics between the SGLT2i group (n=441) and the sulfonylureas group (n=1,420). The absolute standard mean differences (ASMD) for all baseline characteristics were less than 0.1, indicating no substantial difference between the two groups (Figure S1 and Table S6).

**Figure 1.**
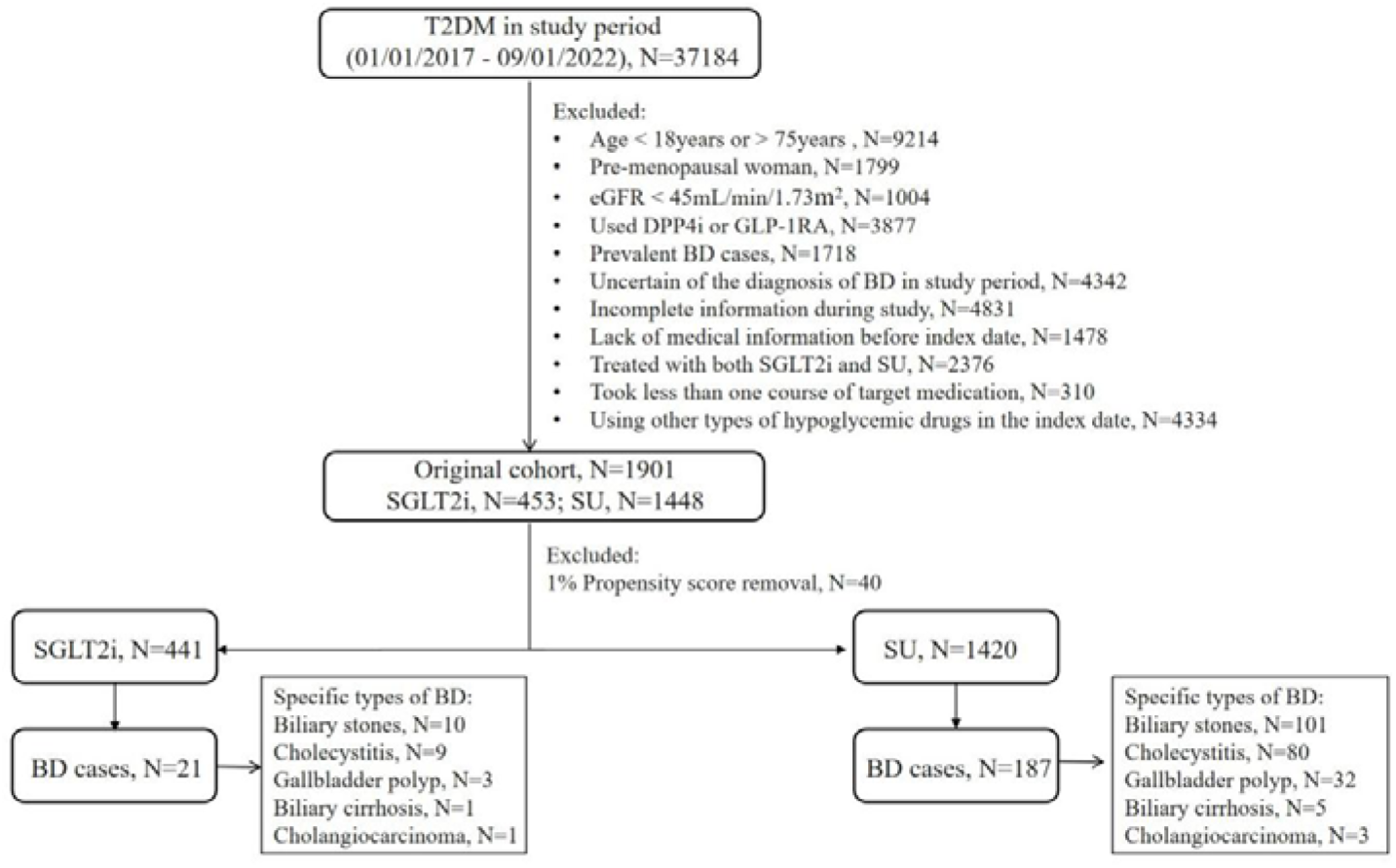
Flowchart of cohort study. Abbreviations: T2DM, type 2 diabetes mellitus; eGFR, estimated glomerular filtration rate; DPP4i, dipeptidyl peptidase-4 inhibitors; GLP-1RA, glucagon like peptide-1 receptor agonists (including canagliflozin, dapagliflozin and empagliflozin); BD, biliary disease (including cholecystitis, gallbladder polyps, and biliary stones); SGLT2i, sodium-glucose cotransporter-2 inhibitors (including canagliflozin, empagliflozin, and dapagliflozin); SU, sulfonylureas (including glipizide, gliclazide, gliquidone, glimepiride, and glyburide); N, number.

### SGLT2i reduced the risk of biliary diseases in patients with T2DM

In the initial unadjusted analysis, patients using SGLT2i exhibited a reduced risk of developing biliary diseases (Hazard Ratio [HR], 0.514; 95% Confidence Interval [CI], 0.311-0.808) in comparison to those starting treatment with sulfonylureas. After excluding the top and bottom 1% of values adjusted for IPTW, the risk remained lower in the SGLT2i group (21 events, 22.7/1000 person-years) relative to the sulfonylurea initiators (187 events, 43.09/1000 person-years), with an HR of 0.527 and 95% CI of 0.319-0.829 (Table 1 and Figure 2A). In the SGLT2i group, 10 patients had cholelithiasis, 9 had cholecystitis, 3 had gallbladder polyps, 1 had bile reflux, and 1 had cholangiocarcinoma. Among them, 3 patients had both cholelithiasis and cholecystitis. In the SU group, 101 patients had cholelithiasis, 80 had cholecystitis, 32 had gallbladder polyps, 5 had bile reflux, and 3 had cholangiocarcinoma. Of these, 34 patients had both cholelithiasis and cholecystitis (Figure 1).

**Figure 2.**
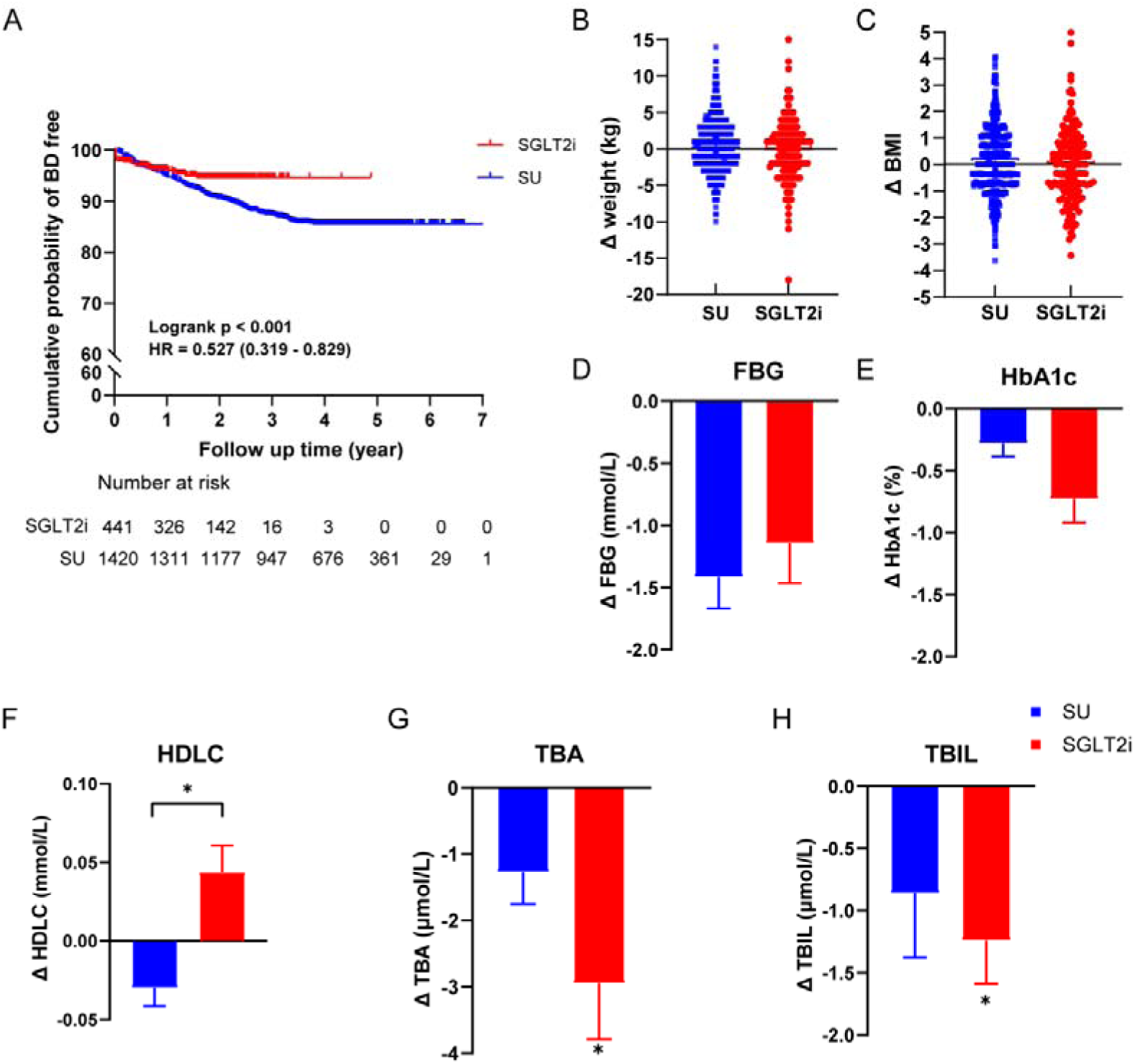
Effect of SGLT2i on the incidence of biliary diseases and clinical index in T2DM. **(A)** SGLT2i significantly reduced the incidence of BS in T2DM. **(B)** There was no change in FBG between the two groups. **(C)** There was no change in HbA1c between the two groups. **(D)** SGLT2i significantly increased the HDLC in T2DM compared with SU treatment. **(E)** TBA level in SGLT2i new users was decreased in T2DM compared to SU new users. **(F)** TBIL level in SGLT2i new users was decreased in T2DM compared to SU new users. All changes were estimated from the index date to the termination date of follow-up. *p<0.05. Abbreviation: SGLT2i, sodium-glucose cotransporter-2 inhibitors; SU, sulfonylureas; HR, hazard ratios; FBG, fasting blood glucose; HbA1c, glycosylated hemoglobinc; HDLC, high density lipoprotein cholesterol; TBA, total bile acid; TBIL, total bilirubin.

**Table 1.** Risk of biliary disease for SGLT2i compared to the SU group.

|  |  | Patients | Biliary disease | Incidence density (per 1000 person-year) | HR (95%CI) |
| --- | --- | --- | --- | --- | --- |
| Crude analysis |  |  |  |  |  |
|  | SGLT2i | 453 | 21 | 21.69 | **0.514 (0.311-0.808)** |
|  | SU | 1448 | 189 | 42.67 | Reference |
| Main analysis |  |  |  |  |  |
|  | SGLT2i | 441 | 21 | 22.70 | **0.527 (0.319-0.829)** |
|  | SU | 1420 | 187 | 43.09 | Reference |
| Outcomes at 24 months |  |  |  |  |  |
|  | SGLT2i | 441 | 21 | 22.70 | **0.616 (0.371-0.974)** |
|  | SU | 1420 | 160 | 36.88 | Reference |
|  | p for trend |  |  |  | **0.028\*** |
| Sensitivity analysis |  |  |  |  |  |
| 1:1 PSM |  |  |  |  |  |
|  | SGLT2i | 430 | 20 | 22.08 | **0.552 (0.311-0.945)** |
|  | SU | 430 | 50 | 39.97 | Reference |
| 1:2 PSM |  |  |  |  |  |
|  | SGLT2i | 430 | 20 | 22.08 | **0.539 (0.314-0.885)** |
|  | SU | 707 | 86 | 40.95 | Reference |
| 1:3 PSM |  |  |  |  |  |
|  | SGLT2i | 430 | 20 | 22.08 | **0.557 (0.327-0.903)** |
|  | SU | 904 | 108 | 39.63 | Reference |
| SMRW after removing 1% |  |  |  |  |  |
|  | SGLT2i | 441 | 21 | 22.70 | **0.527 (0.319-0.829)** |
|  | SU | 1420 | 187 | 43.09 | Reference |
| Logistic regression |  |  |  |  |  |
|  | SGLT2i | 441 | .. | .. | 0.379 (0.234-0.614) |
|  | SU | 1420 | .. | .. | Reference |

Crude analysis was performed on 1901 patients while the main analysis was performed on 1861 patients after 1% removal with IPTW. Abbreviation: SGLT2i, sodium-glucose cotransporter-2 inhibitors; SU, sulfonylureas; HR, hazard ratios; CI, confidence intervals; PSM, propensity score matching; SMRW, standardized mortality ratio weighting; IPTW, inverse probability of treatment weighting.

Multiple sensitivity analyses supported the robustness of these results, including matched analyses using 1:1, 1:2, and 1:3 propensity score matching (PSM) (HRs of 0.552, 0.539, and 0.557, respectively, with corresponding 95% CIs showing statistical significance), standardized mortality ratio weighting (SMRW) after removing the 1% extremes (HR, 0.527; 95% CI, 0.319-0.829), and logistic regression analysis (HR, 0.379; 95% CI, 0.234 - 0.614), all listed in Table 1. Furthermore, when excluding individuals who developed biliary diseases within 24 months post-index date, the results (HR, 0.616; 95% CI, 0.371-0.974) remained consistent with the primary analysis, with a p-value for trend at 0.028 (Table 1 and Table S7).

Figure 2A illustrates the cumulative probability of remaining free from biliary diseases for both groups. Interesting patterns emerged from subgroup analyses, particularly among postmenopausal female patients (HR, 0.190; 95% CI, 0.022-0.724), patients aged over 60 (HR, 0.490; 95% CI, 0.229-0.934), patients without diabetic complications (HR, 0.432; 95% CI, 0.169-0.929), and those with higher aCCI scores (HR, 0.419; 95% CI, 0.204-0.771), as shown in Table S8. Notably, the findings were consistent regardless of the specific type of SGLT2i used.

### No significant differences in weight, BMI, FPG, and HbA1c between SGLT2i Group and SU Group

At baseline, the median (interquartile range) [M (IQR)] of weight and mean BMI of the two groups were comparable, with no statistically significant differences (SGLT2i group: M (IQR) weight = 69 (60, 80) kg, mean BMI=25.30 kg/m^2^; Sulfonylureas group: M (IQR) weight = 69 (64, 76) kg, mean BMI=25.36 kg/m^2^)(Table S9). Throughout the follow-up period, changes in both weight and BMI within each group were minimal and showed no statistically significant differences between the groups. Specifically, by the end of the study, the M (IQR) weight and mean BMI remained similar (SGLT2i group: M (IQR) weight = 69 (61.75,78)kg, mean BMI = 25.28 kg/m^2^; Sulfonylureas group: M (IQR) weight = 70 (64,76) kg, mean BMI = 25.50 kg/m ^2^), indicating that the treatment had no significant impact on these parameters(Figures 2B and 2C). Besides, we analyzed differences in clinical measurements such as fasting plasma glucose (FPG), hemoglobin A1c (HbA1c), and blood biochemical indices between the SGLT2i group and the sulfonylureas group at the start and end of the follow-up period, as presented in Table S9. The initial mean (SD) HbA1c levels were 8.18 (1.78) % for the SGLT2i group and 7.89 (2.10) % for the sulfonylurea group, which by the final follow-up had altered to 7.44 (1.57) % and 7.61 (2.00) %, respectively. Although blood glucose levels decreased from baseline in both groups, there was no significant difference in these changes between the groups (Figures 2D and 2E). This suggests that the effects of SGLT2i on biliary diseases may not be related to their hypoglycemic effect.

### Significant reduction in TBA and TBIL and increase in HDLC observed in SGLT2i Group

Regarding blood lipids, both groups showed a significant reduction in total cholesterol (TC) (SGLT2i group from 4.41 (3.70, 5.16) to 4.32 (3.53, 5.27), p=0.021, and the sulfonylureas group from 4.41 (3.76, 5.19) to 4.29 (3.59, 5.09), p<0.001) and triglycerides (TG) (SGLT2i group from 1.58 (1.09, 2.46) to 1.43 (1.05, 1.93), p<0.001 and the sulfonylurea group from 1.52 (1.06, 2.21) to 1.44 (1.02, 2.03), p<0.001), with no significant differences in the magnitude of these changes between the groups. However, a notable increase in HDLC (p = 0.002) occurred in the SGLT2i group by the study’s end. Additionally, serum TBA (p < 0.001) and TBIL (p = 0.014) levels significantly decreased in the SGLT2i group compared with the sulfonylurea group (Figures 2F-2H and Table S9).

### SGLT2i reduced the incidence of gallstones and improved gallbladder emptying function in LD-fed mice

All groups of mice fed on a lithogenic diet (LD) consumed comparable amounts of food and water and exhibited similar weight gain after eight weeks, as documented in Figure S2A-C. During the 8 weeks of LD and normal diet (ND) feeding, we monitored the occurrence of gallstone formation. The livers of the ND+placebo and LD+henagliflozin groups had a healthier, redder appearance, displayed similar reductions in the liver index, and the bile in their gallbladders appeared clearer (Figure S2D and Figure 3A).

**Figure 3.**
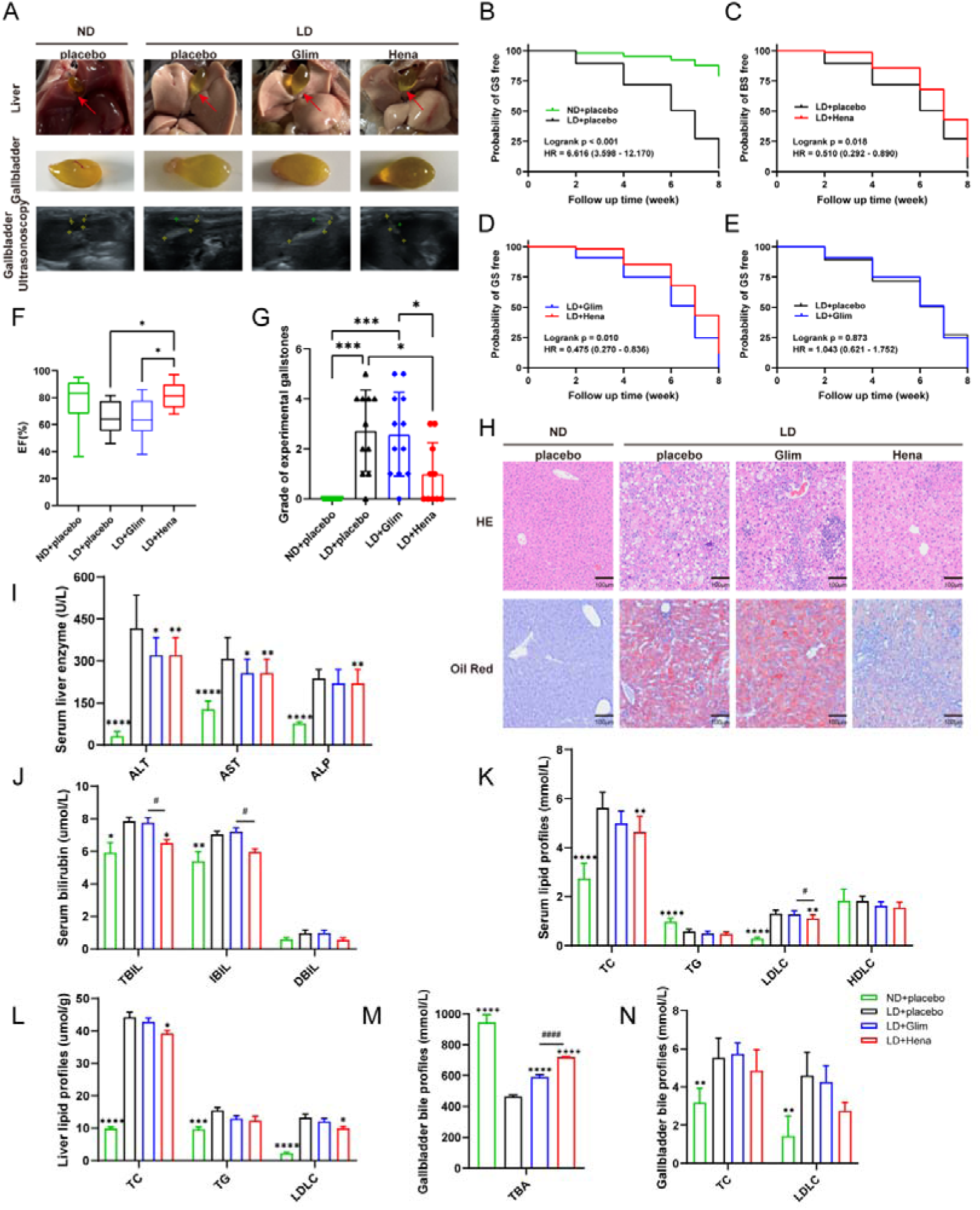
The incidence of gallstones, changes in gallbladder emptying function and liver function in LD-fed mice and ND-fed mice after 8 weeks of treatment. **(A)** Representative images of the mice livers and gallbladders. **(B)** Probability of GS free in ND mice and LD mice after 8 weeks of treatment. **(C)** Probability of GS free in LD+Hena mice and LD+placebo mice after 8 weeks of treatment. **(D)** Probability of GS free in LD+Hena mice and LD+Glim after 8 weeks of treatment. **(E)** Probability of GS free in LD+Glim mice and LD+placebo after 8 weeks of treatment. **(F)** Gallbladder emptying function in different groups. **(G)** Grade of experimental gallstones. **(H)** Liver HE staining and oil red staining of mice liver. All magnifications are ×200. **(I)** Serum liver function in mice. **(J)** Serum bilirubin in mice. **(K)** Serum lipid profiles in mice. **(L)** Lipid profiles in mice liver. **(M)** TBA in mice gallbladder bile. **(N)** Lipids in mice gallbladder bile. *The asterisk indicates a statistically significant difference between other groups and LD-fed with placebo mice; #The pound key indicates a statistically significant difference between LD-fed with henagliflozin mice and LD-fed with glimepiride treatment mice; *p<0.05, **p<0.01, ***p<0.001, ****p<0.0001. ^#^p<0.05, ^##^p<0.01, ^###^p<0.001, ^####^p<0.0001.

After 8 weeks of feeding, the incidence of gallstones was 72.7% (8/11) in the LD+henagliflozin group, 100% (12/12) in the LD+glimepiride group, 91.7% (11/12) in the LD+placebo group, and 10% (1/10) in the ND+placebo group (Figure S2E). Furthermore, survival analysis indicated that mice on the LD were 6.6 times more likely to develop gallstones than those on the ND (Figure 3B). Treatment with SGLT2i significantly reduced the occurrence of biliary diseases, with a hazard ratio (HR) of 0.510 and a 95% confidence interval (CI) of 0.292-0.890, compared to placebo (Figure 3C). Additionally, SGLT2i treatment led to a notable 52% reduction in gallstone incidence compared to sulfonylureas treatment in LD-fed mice (Figure 3D). In contrast, sulfonylureas treatment did not produce such an effect (Figure 3E). Most importantly, SGLT2i notably improved gallbladder emptying function. The value of ejection fraction (EF) was 82.52±2.97% in LD-fed mice treated with henagliflozin, compared to 64.70±4.01% with glimepiride treatment, 65.50±3.07% with placebo, and 77.11±6.62% in ND-fed mice (Figure 3F). In line with this finding, the expression levels of the Cholecystokinin (Cck) and Gastrin genes were increased in the livers of mice treated with henagliflozin (Figure 5C). It is well established that cholecystokinin and gastrin are the primary gut hormones secreted after a meal, responsible for regulating gastric acid secretion, digestive enzymes, and gallbladder emptying.^22^ Moreover, in terms of the severity of experimental gallstones, mice treated with henagliflozin had lower grades compared with those treated with glimepiride or placebo (Figure 3G and Table S10).

### SGLT2i alleviated liver injury and dyslipidemia in the LD-fed mice model

Given that the development of cholelithiasis is associated with obesity, hyperlipidemia, non-alcoholic fatty liver disease, and other conditions^23, 24^, we investigated a range of contributing factors. In addition to gallbladder dysmotility, gallstone formation is also linked to genetic factors, excessive hepatic cholesterol secretion (leading to cholesterol supersaturation in gallbladder bile), and the accelerated growth and solidification of cholesterol crystals.^25^ We, therefore, examined liver tissue and serum and bile biochemistry to determine whether treatment with henagliflozin affects these associated risk factors. HE staining of liver sections revealed that mice in the LD+placebo and LD+glimepiride groups exhibited clear vacuolar degeneration and edema, along with significant inflammatory cell infiltration, including neutrophils, lymphocytes, and plasma cells, compared to the ND group (Figure 3H). Oil Red O staining showed marked lipid accumulation in the livers of mice treated with placebo or glimepiride on the LD. In contrast, mice treated with henagliflozin on the LD showed significantly less lipid accumulation, reduced inflammatory cell infiltration, and livers more closely resembling those on an ND (Figure 3H).

Biochemical analysis of serum and bile indicated that the LD led to impaired liver function and dyslipidemia in mice, as evidenced by increased levels of ALT (487.00±268.39), AST (300.50±105.65), and ALP (237.17±32.33) compared to the ND. These markers of liver injury were notably reduced following henagliflozin treatment (Figure 3I and Table S11). Henagliflozin treatment also resulted in lower serum levels of total TC and LDLC under fasting conditions in serum, bile, and liver tissue (Figure 3K, 3L, 3N, and Table S10). Since excessive bilirubin is also implicated in gallstone formation, we measured bilirubin levels in bile and serum, finding that LD feeding significantly elevated serum TBIL, IBIL, and DBIL levels compared with the ND. Henagliflozin treatment mitigated this elevation (Figure 3J and Table S11). Furthermore, considering the link between blood glucose, insulin resistance, and cholelithiasis, we measured FBG and serum insulin levels across the groups to ascertain whether the impact of henagliflozin on gallstone formation is independent of blood glucose-related factors. Interestingly, no differences were observed in FBG, random blood glucose (RBG), FINS, and HOMA-IR before and after treatment across the four groups (Table S11 and Figure S2F-I).

These results suggest that henagliflozin ameliorates liver injury and dyslipidemia in the gallstone mice model, thereby reducing the risk of gallstone formation independently of its effects on blood glucose.

### SGLT2i reduced 12α-OH bile acids and promoted primary bile acid biosynthesis and bile acids excretion from feces in the LD-fed mice model

To elucidate how SGLT2i may influence bile acid metabolism and affect gallstone formation, we conducted metabolomic profiling of ileum contents and serum samples to identify specific bile acid components that could impact host metabolism. Despite no significant differences in the principal component analysis (PCA) scores among the LD-fed mice receiving the three different treatments (Figure 4A), there were notable changes in the bile acid composition. Henagliflozin treatment primarily increased TBA in gallbladder bile (Figure 3L) and diminished TBA in serum and ileal contents (Figure 4B and S3B), but had no effect on colon contents (Figure S4B). This was particularly evident in the reduction of 12α-hydroxylated (12α-OH) bile acids in mice treated with henagliflozin, such as β-Muricholic acid (β-MCA), Murideoxycholic acid (MDCA), Deoxycholic acid (DCA), Cholic acid (CA), Allocholic acid (ACA), Glycodeoxycholic Acid (GDCA), Taurodeoxycholic Acid (TDCA), and Taurocholic acid (TCA) (Figure 4C and 4D, Table S12). An increase in 12α-OH bile acids has been associated with metabolic diseases like diabetes and obesity^26^ and reducing the ratio of 12α-OH to non-12α-OH bile acids may improve glycemic and lipid metabolism.^27^ Furthermore, KEGG pathway analysis linked these bile acid deviations primarily to cholesterol metabolism, primary bile acid biosynthesis, and taurine and hypotaurine metabolism (Figure 4E and 4F, Table S13). The modifications in bile acid metabolism observed in ileal contents paralleled these findings (Figure S3), while bile acid metabolism in colon contents had no significant difference in these three groups (Figure S4). These data suggest that henagliflozin treatment can partially normalize bile acid homeostasis in the biliary fluid of the LD-fed mice model.

**Figure 4.**
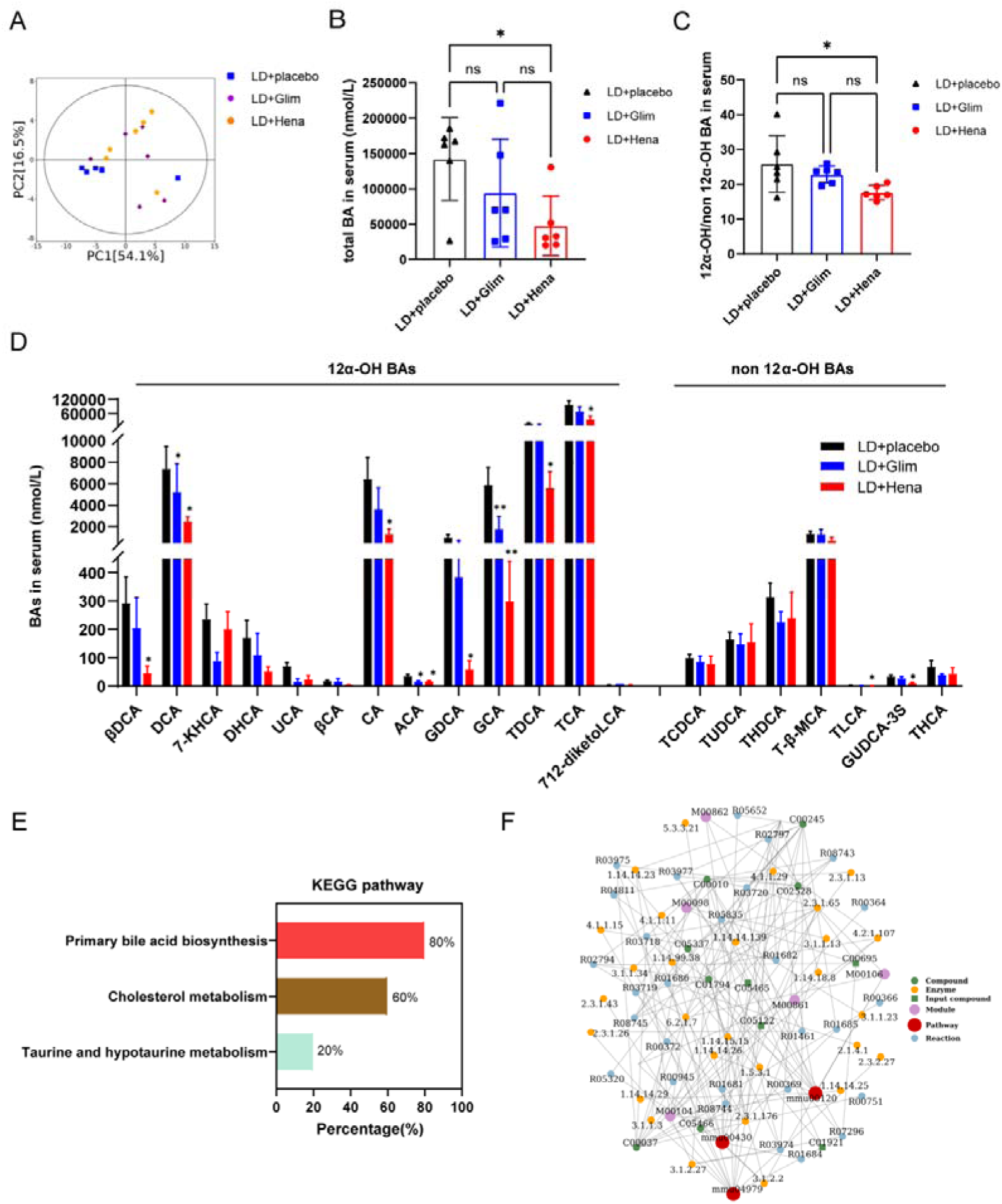
Serum bile acid metabolomics analysis. **(A)** Principal component analysis (PCA) in three different LD-fed mice groups after eight weeks of treatment. **(B)** Total BA in serum. **(C)** 12α-OH/non 12α-OH BAs in serum. **(D)** Relative quantification of specific bile acid components. **(E)** KEGG analysis. **(F)** Network analysis for three groups. The red dots represent a metabolic pathway, and the yellow dots represent relevant regulatory enzyme information. *The asterisk indicates a statistically significant difference between LD-fed with henagliflozin or glimepiride treatment and LD-fed with placebo mice.*p<0.05, **p<0.01.

### SGLT2i modulated the expression of genes linked to cholesterol metabolism

In vitro results showed that treatment with the SGLT2 inhibitor T-1095 increased the expression of the hepatic canalicular cholesterol transporters ATP-binding cassette subfamily G members 5 and 8 (Abcg5/8) in primary mouse and human hepatocytes (Figure 5A). Similarly, the *Ldlr* gene, which is involved in cholesterol uptake and efflux, was also upregulated in both liver tissue and hepatocytes (Figures 5A, 5D, and 5F). However, no significant difference in the expression of *Abcg5/8* was observed in the livers of LD-fed mice treated with henagliflozin compared to LD-fed mice without treatment. These findings indicated that SGLT2i exerts an influence on cholesterol metabolism, which in turn impacts lipid profiles.

**Figure 5.**
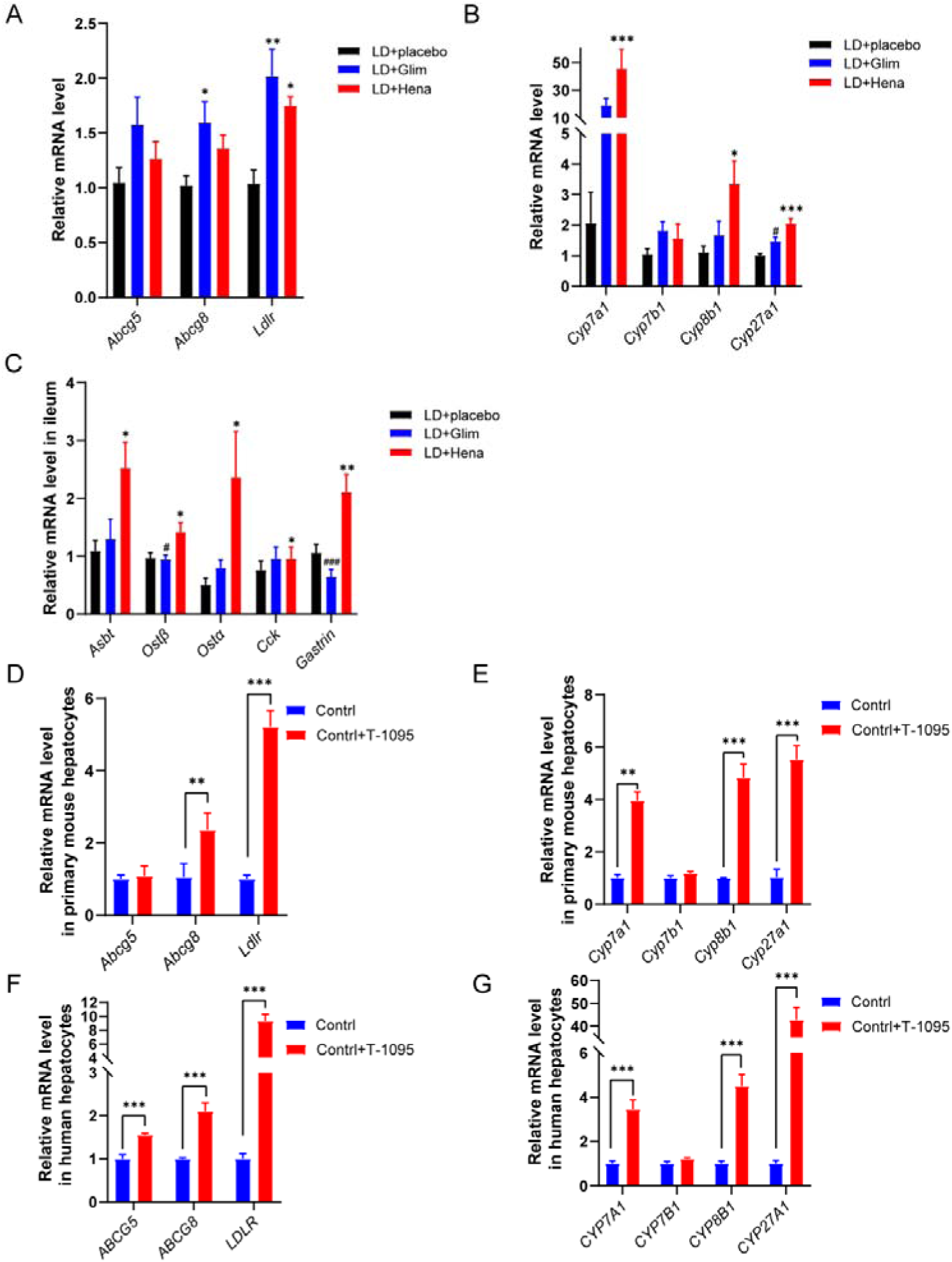
Validation in mouse liver and ileum, primary mouse hepatocytes and human hepatocytes. **(A)** Expression of mRNA levels of lipid metabolism-related genes in the liver. **(B)** Expression of mRNA levels of bile acid metabolism-related genes in liver. **(C)** Expression of mRNA levels of bile acid metabolism-related genes and genes involved in gallbladder contraction in ileum. **(D)** Expression of mRNA levels of lipid metabolism-related genes in primary mouse hepatocytes after 10μM T-1095 treated for 24h. **(E)** Expression of mRNA levels of bile acid metabolism-related genes in primary mouse hepatocytes treated with T-1095 for 24h. **(F)** Expression of mRNA levels of lipid metabolism-related genes in human hepatocytes after 10mM T-1095 treated for 24h. **(G)** Expression of mRNA levels of bile acid metabolism-related genes in human hepatocytes treated with T-1095. *The asterisk indicates a statistically significant difference between LD-fed with henagliflozin mice and LD-fed with glimepiride treatment mice. *p<0.05, **p<0.01, ***p<0.001, ****p<0.0001. ^#^The pound key indicates a statistically significant difference between LD-fed with henagliflozin mice and LD-fed with glimepiride treatment mice. ^#^p<0.05, ^##^p<0.01, ^###^p<0.001, ^####^p<0.0001.

### SGLT2i modulated the expression of genes associated with bile acid metabolism

Bile acids (BAs) are a class of steroid acids produced in the liver from cholesterol through several enzymatic reactions along two primary pathways. Approximately 75% of BA synthesis occurs through the main pathway, also known as the classical or neutral pathway, which begins with the 7α-hydroxylation of cholesterol by the enzyme CYP7A1. This is followed by further modification of the steroid nucleus and the oxidative cleavage of the side chain, a step in which the enzyme CYP8B1 is involved. The alternative pathway starts with the 27-hydroxylation of cholesterol by CYP27A1. The resulting oxysterols are further hydroxylated by oxysterol 7α-hydroxylase (CYP7B1). The majority of bile acids (about 95%) are then reabsorbed in the ileum and returned to the liver via enterohepatic circulation, while the remaining 5%, which are not reabsorbed, are excreted in feces.^28^ Synthesis and excretion of BAs are major routes of cholesterol and lipid catabolism. To maintain bile acid pool homeostasis, the hepatic synthesis of BAs must balance the amount lost through fecal excretion. As such, impeding BA reabsorption enhances fecal BA excretion, stimulating de novo synthesis of BAs from cholesterol and potentially countering obesity induced by a high-fat diet.^29^

Examining the expression of BA metabolism-related genes in mouse liver revealed significant upregulation of *Cyp7a1*, *Cyp8b1*, and *Cyp27a1* (Figures 5B, 5E, and 5G), which suggests that the capacity for BA synthesis was considerably increased following treatment with henagliflozin. This may be attributable to heightened bile acid excretion. Other essential proteins include the apical sodium-dependent bile acid transporter (ASBT, SLC10A2), which mediates the entry of bile acids into enterocytes. Organic solute transporters alpha and beta (OSTα, OSTβ) are believed to be the primary bile acid efflux transporters in mammalian intestines and thus are vital for bile acid homeostasis and enterohepatic circulation.^30^ Our findings indicated that the expression of *Asbt*, *Ostα*, and *Ostβ* in ileal tissue was upregulated after henagliflozin treatment (Figure 5C). According to the results in Figure S4, henagliflozin treatment could eliminate almost the same amount of bile acids from the body as other treatments, these results suggest that henagliflozin inhibits the reabsorption of bile acids and promotes the excretion of bile acids from feces, thereby maintaining bile acid homeostasis in response to a reduction in serum bile acids.

## Discussion

Here, we present data from a retrospective cohort study indicating that the use of SGLT2i was associated with a 38% lower risk of biliary diseases (HR 0.595, 95% CI 0.410-0.863) in patients with T2DM compared to those using sulfonylureas. This effect is particularly significant in specific patient subgroups, including females (HR 0.190, 95% CI 0.022-0.724), individuals aged over 60 years (HR 0.490, 95% CI 0.229-0.934), those without diabetic complications (HR 0.432, 95% CI 0.169-0.929), and those with more clinical comorbidities (HR 0.419, 95% CI 0.204-0.774). These effects were observed irrespective of the specific types of SGLT2i used. Furthermore, the beneficial impact on reducing the risk of biliary diseases appears to correlate with the duration of SGLT2i treatment particularly when treatment extends beyond 24 months. Consistent with the clinical findings, SGLT2i administration markedly prevents gallstone formation in the mice fed with a lithogenic diet. Mechanistically, SGLT2i makes this beneficial impact by lowering the cholesterol level in the blood and reducing the ratio of 12α-OH/non 12α-OH bile acids, therefore potentially ameliorating the supersaturation in gallbladder bile as well as enhancing the emptying ability of the gallbladder. Our findings support a novel mechanism of SGLT2i in the prevention of cholelithiasis, particularly beyond their glycemic effects.

It was reported that the prevalence and incidence of gallstones were 5%-25%, and 0.47 per 100 person-years, respectively. Diabetes has been thought one of the major risk factors for gallstones. About 33.3% of people with diagnosed diabetes had gallbladder diseases (RR 2.3 [1.7-3.2] compared with no diabetes); including 15.4% with ultrasound-diagnosed gallstones (RR 1.6 [1.1-2.4]) and 17.9% (RR 2.6 [1.7-4.0]) with cholecystectomy.^3^ A meta-analysis of ten prospective cohort studies indicated the relative risk of gallbladder disease among people with diabetes was 1.56 (1.26-1.93) compared with people without diabetes.^31^ BD can lead to severe complications, including bile duct obstruction, infection, and pancreatitis, and these complications can exacerbate comorbidities like cardiovascular disease and renal impairment, worsening outcomes in T2DM patients. Furthermore, surgical procedures such as cholecystectomy pose higher risks for T2DM patients due to delayed wound healing and increased infection rates. Therefore, the prevention of BD is paramount. However, despite the hypoglycemic drugs used, many patients who reach the treatment target of glycemic control still develop gallstones.^6^ The data from a population-based cohort study reveals an inverse relationship exists between type 1 diabetes and gallstones in patients aged twenty to forty years although a strong association between T2DM and gallstone diseases was found.^32^ These findings suggested that hyperglycemia may not be the independent risk factor for cholelithiasis. In the current study, we followed up 1,901 diabetic patients who have no biliary diseases at the start time point of treatment until the earliest onset of any biliary diseases. Meanwhile, the patient selection in our study excluded the ones accepted treatment of incretin-based drugs (GLP-1RA and DPP4 inhibitors) given they were associated with an increased risk of biliary diseases.^6^ Additionally, differences in the risk of biliary disorders were noticed as early as twenty-four months following the start of sulfonylureas or SGLT2i medication.

By the way, we chose sulfonylureas as the control group due to their common use in similar studies,^33, 34^, established baseline for comparison, overlapping treatment guidelines, comparable baseline characteristics, and controlled confounding factors. Remarkably, there was a rare occurrence of biliary diseases among patients treated with SGLT2i. The benefits of SGLT2i in reducing the risks of biliary diseases, as observed in the current cohort study, could offer valuable insights for choosing glucose-lowering treatments in patients with T2DM, especially female and senior people who face a risk of developing biliary diseases, including cholelithiasis.

While enhanced glycemic control has traditionally been deemed crucial in mitigating the risk of hyperglycemia and its associated complications, as a result of the use of sulfonylureas as an active comparator arm in this study, the differences in patients’ weight, blood glucose, and HbA1c between the two groups during the trial were modest and no significant disparity in these changes was noted between the two groups. Consequently, these findings indicate that the benefits of the improvement of biliary diseases are unlikely mediated by the glucose-lowering and weight-lowering properties of SGLT2i.

Although cholesterol cholelithiasis has been regarded as a multifactorial disease influenced by a complex interaction of genetic and environmental factors, the contribution of blood glucose to gallstone formation still remains controversial.^32^ In our study, given all the patients treated with either SGLT2i or sulfonylureas reached the target of glycemic control, the incidence of cholelithiasis was markedly different. This data indicates hyperglycemia may not be an independent risk factor of cholelithiasis and the preventive effect of SGLT2i may be beyond glycemic control. In terms of pathogenesis, cholesterol supersaturation and gallbladder hypomotility are essential components that occur in concert during the initiation and propagation of cholesterol stones, which represent more than 90% of biliary stones.^25^ It has been thought lowering hepatic cholesterol content decreases its bioavailability for biliary secretion. Indeed, we observed that SGLT2i induces a significant reduction in the cholesterol level in both the serum and liver accompanied by improved lipid accumulation and hepatic function in mice. In our study, the lipid compositions of gallbladder bile from mice treated with the SGLT2i show a noticeable decrease in cholesterol saturation and LDLC level in bile. Considering their role in promoting cholesterol elimination from the body, bile acid synthesis serves as an indicator of cholesterol turnover in the liver. Treatment with SGLT2i significantly decreased total bile acid levels, particularly 12α-OH bile acids in the mice serum and ileum contents and increased the total bile acids excretion from the feces. These 12α-OH bile acids have been associated with gallstone formation due to their diminished capacity to solubilize cholesterol compared to other bile acids.^35^ The dysfunction of gallbladder motility typically results in the paralysis of its contractile function, consequently impairing its ability to empty properly. This extended stasis creates an environment conducive to the nucleation of cholesterol crystals and their subsequent aggregation into macroscopic stones.^36^ Collectively, the mitigated lithogenic effects of SGLT2i are primarily due to the reduction in biliary cholesterol content within the bile and enhanced gallbladder motility induced by CCK and gastrin from the ileum.

To the best of our knowledge, the current study is the first clinical investigation involving a multicenter retrospective cohort to compare the risk of developing biliary diseases with SGLT2i and sulfonylurea treatment. As one of its advantages, our cohort study with active comparator new-users was designed to minimize selection bias and exclude numerous relevant confounding factors, resulting in a more representative selected population. Moreover, we repeatedly validated our results through multiple sensitivity analyses, including the application of propensity score overlap weighting, which closely aligned with our main findings, thereby enhancing their credibility. To further validate the clinical findings, we also conducted animal experiments to elucidate the preventive mechanism of SGLT2i on the risk of gallstone formation in lithogenic diet-fed mice. The findings of this study offer a new potential application of SGLT2i.

However, there are still several limitations in this study. Because this is a retrospective study, we lacked certain testing parameters such as patient serum insulin levels. Consequently, we were unable to determine definitively whether SGLT2i reduces the incidence of biliary diseases by improving insulin resistance. However, these limitations were addressed by the subsequent animal experiment. Furthermore, this study only provides a preliminary exploration of the specific mechanisms underlying the association between SGLT2i and a reduced risk of gallstones. Further detailed validation is necessary. Therefore, additional prospective clinical and basic research is required to investigate the relationship between SGLT2i and biliary diseases.

## Conclusion

We conducted a multicenter retrospective cohort analysis to establish a novel association between the use of SGLT2i and a lower risk of biliary diseases, particularly in postmenopausal women, those aged 60 and above, those without diabetes complications, and those with a higher prevalence of clinical comorbidities in clinical practice. Mechanistically, the protection of SGLT2i on cholelithiasis potentially may result from their effect on improved bile metabolism and gallbladder motility. Our findings offer an potential application of SGLT2i beyond glucose control in the future.

## Data Availability

All data produced in the present study are available upon reasonable request to the authors

## Author contributions

Ming Gao: conceptualization, formal analysis, methodology, software, validation, writing-original draft. Qiuyu Lin: data curation, methodology. Tingyi Zhu: data curation and visualization. Kaiyue Hu: data curation and methodology. Bei Zhong: methodology. Kaini Zhang: methodology and visualization. Xiaoli Chen: data couration, methodology. Xinyu Chen and Ying Zhang: data curation. Yangyang Li: supervision. Shaowen Tang: formal analysis, data curation, supervision. Dongming Su: supervision, validation, writing-revise and editing. Xiubin Liang: conceptualization, writing-review and editing. Yu Liu: funding acquisition, writing-review and editing.

## Data Transparency Statement

Data, analytic methods, and study materials will not be made available to other researchers.

Author names in bold designate shared co-first authorship.

---

**Figure S1.**
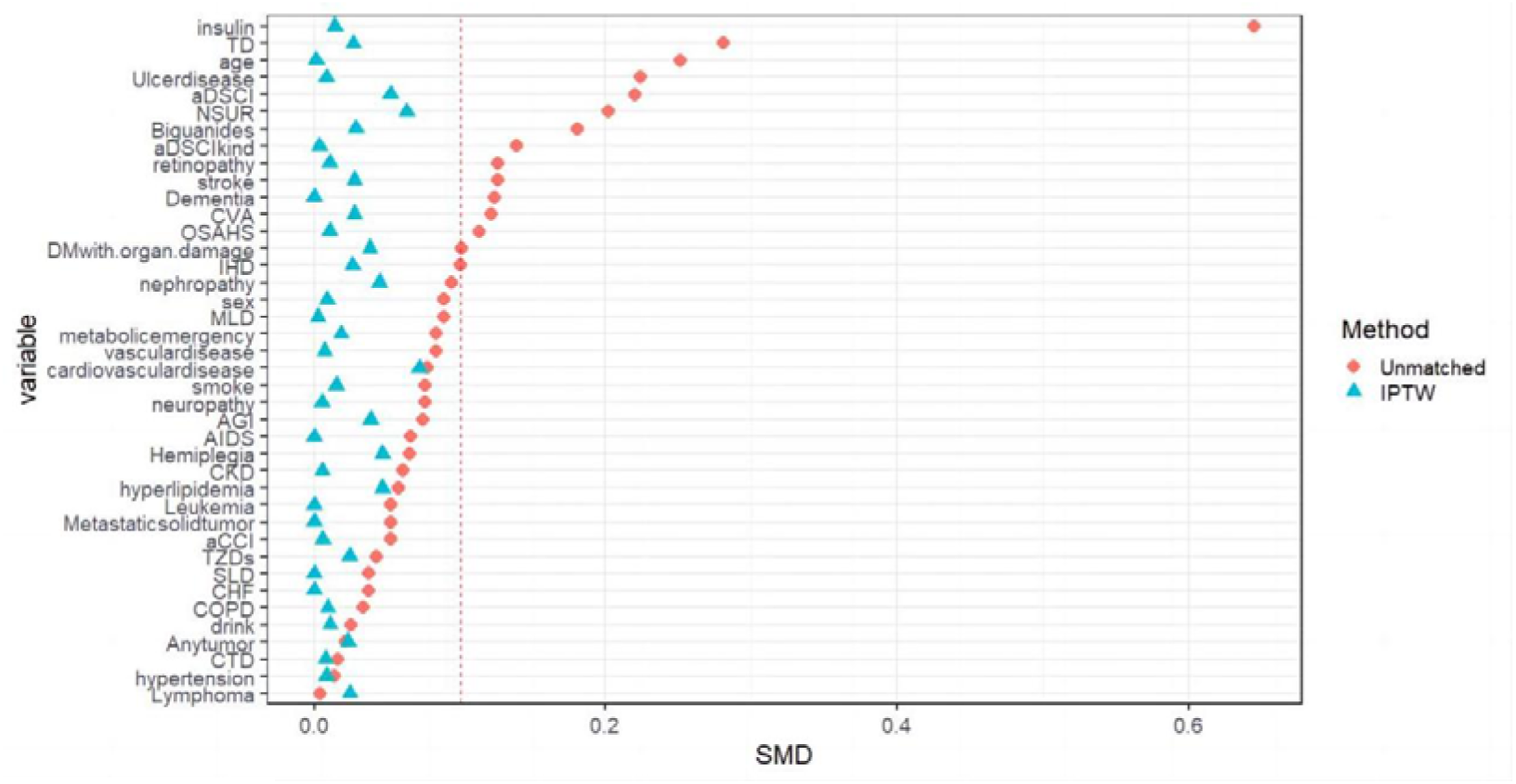

**Figure S2.**
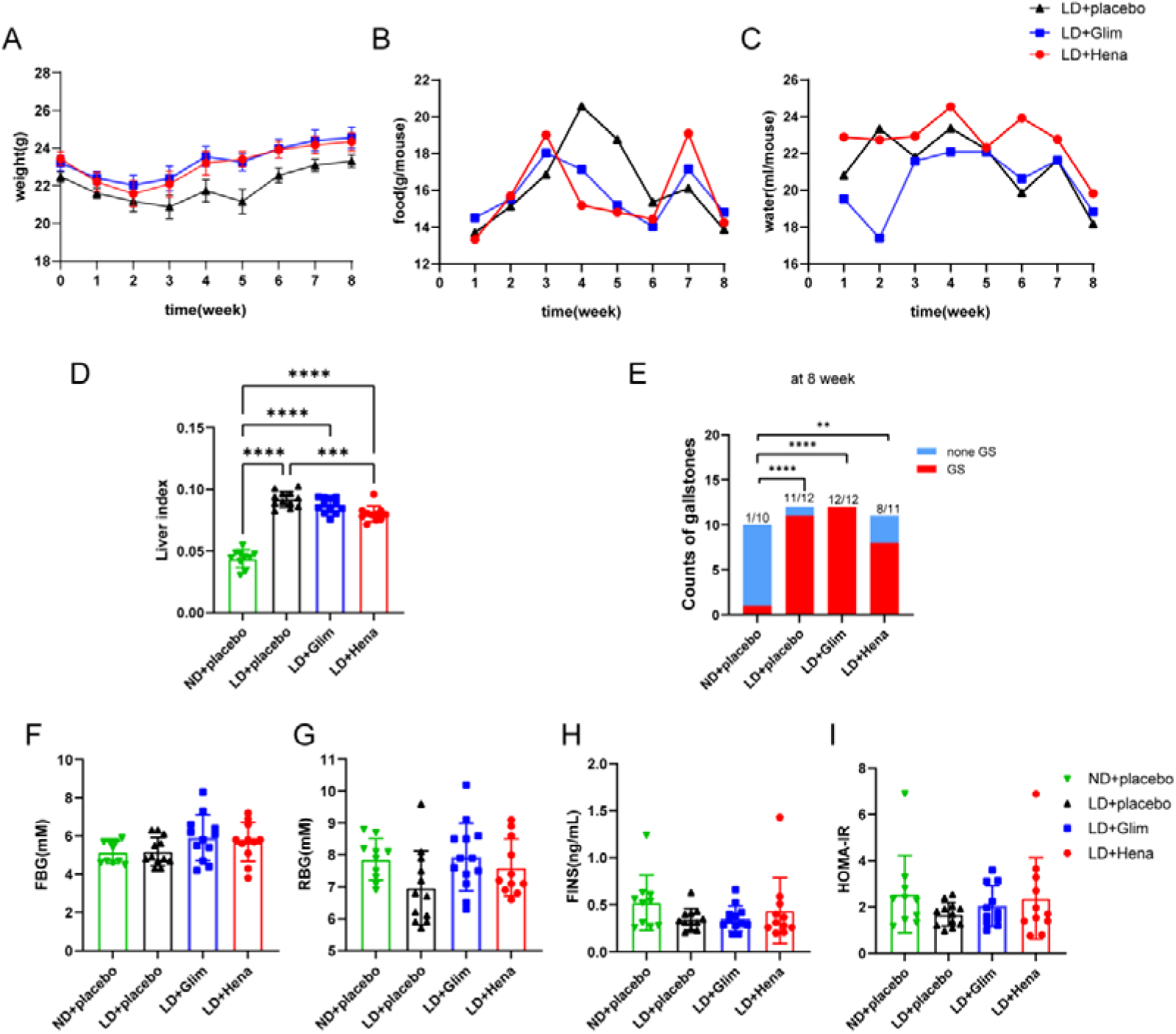

**Figure S3.**
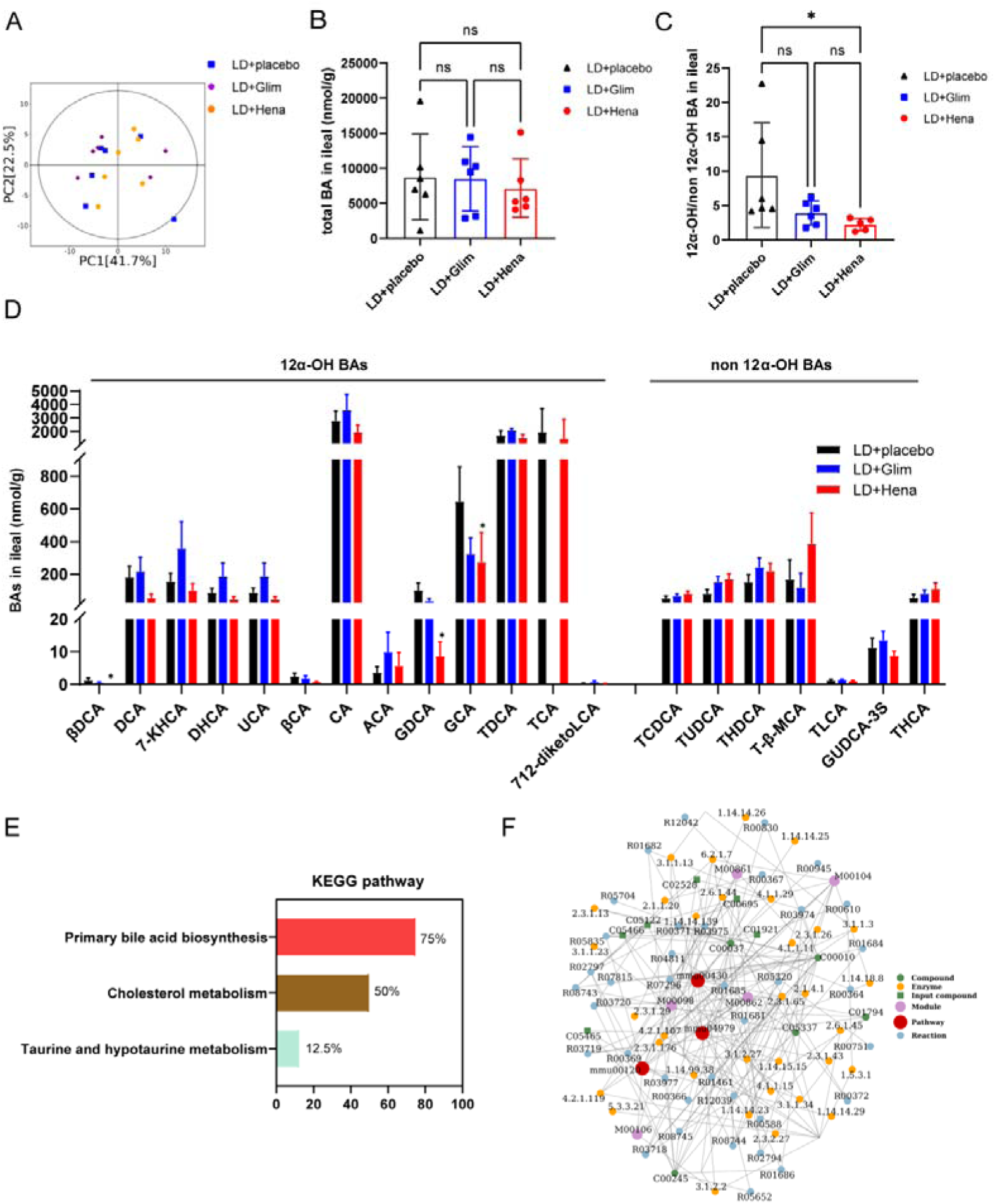

**Figure S4.**
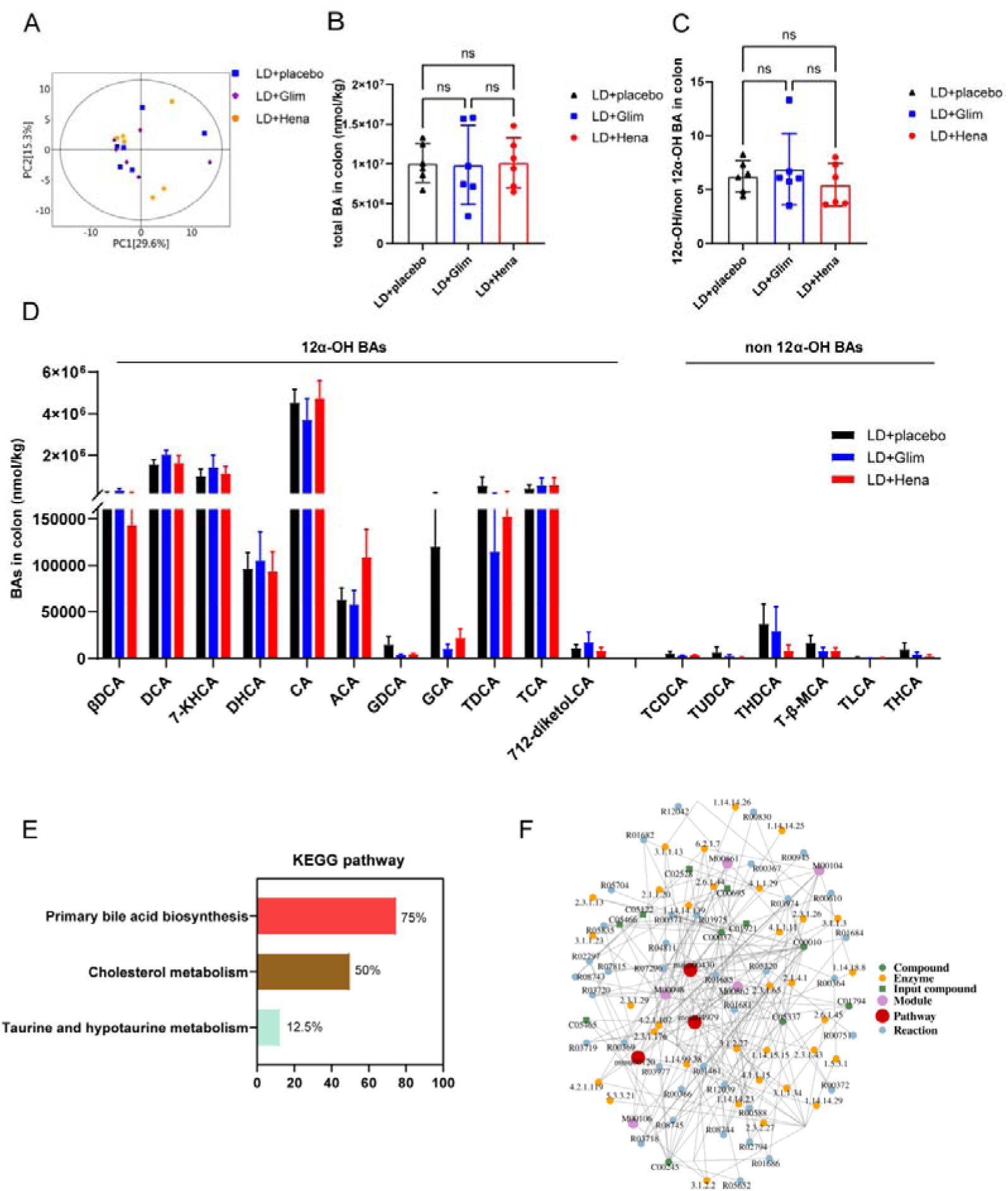

## References

1. Lammert F, Gurusamy K, Ko CW, et al. Gallstones. Nat Rev Dis Primers 2016;2:16024.

2. Wang X, Yu W, Jiang G, et al. Global Epidemiology of Gallstones in the 21st Century: A Systematic Review and Meta-Analysis. Clin Gastroenterol Hepatol 2024.

3. Ruhl CE, Clark JM, Everhart JE. Liver and Gallbladder Disease in Diabetes. In: Cowie CC, Casagrande SS, Menke A, Cissell MA, Eberhardt MS, Meigs JB, Gregg EW, Knowler WC, Barrett-Connor E, Becker DJ, Brancati FL, Boyko EJ, Herman WH, Howard BV, Narayan KMV, Rewers M, Fradkin JE, eds. Diabetes in America. 3rd ed. Bethesda (MD) interest., 2018.

4. Noel RA, Braun DK, Patterson RE, et al. Increased risk of acute pancreatitis and biliary disease observed in patients with type 2 diabetes: a retrospective cohort study. Diabetes Care 2009;32:834–8.

5. He L, Wang J, Ping F, et al. Dipeptidyl peptidase-4 inhibitors and gallbladder or biliary disease in type 2 diabetes: systematic review and pairwise and network meta-analysis of randomised controlled trials. BMJ 2022;377:e068882.

6. Faillie JL, Yu OH, Yin H, et al. Association of Bile Duct and Gallbladder Diseases With the Use of Incretin-Based Drugs in Patients With Type 2 Diabetes Mellitus. JAMA Intern Med 2016;176:1474–1481.

7. Ratheesh R, Ulrich MT, Ghozy S, et al. The association between diabetes and gallstones: a nationwide population-based cohort study. Prz Gastroenterol 2023;18:292–299.

8. Hu J, Teng J, Hui S, et al. SGLT-2 inhibitors as novel treatments of multiple organ fibrosis. Heliyon 2024;10:e29486.

9. Parving HH, Lambers-Heerspink H, de Zeeuw D. Empagliflozin and Progression of Kidney Disease in Type 2 Diabetes. N Engl J Med 2016;375:1800–1.

10. Perkovic V, Jardine MJ, Neal B, et al. Canagliflozin and Renal Outcomes in Type 2 Diabetes and Nephropathy. N Engl J Med 2019;380:2295–2306.

11. Fernandez-Balsells MM, Sojo-Vega L, Ricart-Engel W. Canagliflozin and Cardiovascular and Renal Events in Type 2 Diabetes. N Engl J Med 2017;377:2098.

12. Paik JM, Tesfaye H, Curhan GC, et al. Sodium-Glucose Cotransporter 2 Inhibitors and Nephrolithiasis Risk in Patients With Type 2 Diabetes. JAMA Intern Med 2024;184:265–274.

13. Chang HY, Weiner JP, Richards TM, et al. Validating the adapted Diabetes Complications Severity Index in claims data. Am J Manag Care 2012;18:721–6.

14. Charlson ME, Pompei P, Ales KL, et al. A new method of classifying prognostic comorbidity in longitudinal studies: development and validation. J Chronic Dis 1987;40:373–83.

15. Woods SE, Leonard MR, Hayden JA, et al. Impaired cholecystokinin-induced gallbladder emptying incriminated in spontaneous “black” pigment gallstone formation in germfree Swiss Webster mice. Am J Physiol Gastrointest Liver Physiol 2015;308:G335–49.

16. Akiyoshi T, Uchida K, Takase H, et al. Cholesterol gallstones in alloxan-diabetic mice. J Lipid Res 1986;27:915–24.

17. Li K, Zhang K, Wang H, et al. Hrd1-mediated ACLY ubiquitination alleviate NAFLD in db/db mice. Metabolism 2021;114:154349.

18. Haukoos JS, Lewis RJ. The Propensity Score. JAMA 2015;314:1637–8.

19. Robins JM, Hernan MA, Brumback B. Marginal structural models and causal inference in epidemiology. Epidemiology 2000;11:550–60.

20. Brookhart MA, Wyss R, Layton JB, et al. Propensity score methods for confounding control in nonexperimental research. Circ Cardiovasc Qual Outcomes 2013;6:604–11.

21. Stuart EA, Lee BK, Leacy FP. Prognostic score-based balance measures can be a useful diagnostic for propensity score methods in comparative effectiveness research. J Clin Epidemiol 2013;66:S84–S90 e1.

22. Rehfeld JF. Incretin physiology beyond glucagon-like peptide 1 and glucose-dependent insulinotropic polypeptide: cholecystokinin and gastrin peptides. Acta Physiol (Oxf) 2011;201:405–11.

23. Sun H, Warren J, Yip J, et al. Factors Influencing Gallstone Formation: A Review of the Literature. Biomolecules 2022;12.

24. Andreotti G, Chen J, Gao YT, et al. Serum lipid levels and the risk of biliary tract cancers and biliary stones: A population-based study in China. Int J Cancer 2008;122:2322–9.

25. van Erpecum KJ. Biliary lipids, water and cholesterol gallstones. Biol Cell 2005;97:815–22.

26. Jia W, Wei M, Rajani C, et al. Targeting the alternative bile acid synthetic pathway for metabolic diseases. Protein Cell 2021;12:411–425.

27. Li P, Ruan X, Yang L, et al. A liver-enriched long non-coding RNA, lncLSTR, regulates systemic lipid metabolism in mice. Cell Metab 2015;21:455–67.

28. Schaap FG, Trauner M, Jansen PL. Bile acid receptors as targets for drug development. Nat Rev Gastroenterol Hepatol 2014;11:55–67.

29. Rao A, Kosters A, Mells JE, et al. Inhibition of ileal bile acid uptake protects against nonalcoholic fatty liver disease in high-fat diet-fed mice. Sci Transl Med 2016;8:357ra122.

30. Soroka CJ, Ballatori N, Boyer JL. Organic solute transporter, OSTalpha-OSTbeta: its role in bile acid transport and cholestasis. Semin Liver Dis 2010;30:178–85.

31. Aune D, Vatten LJ. Diabetes mellitus and the risk of gallbladder disease: A systematic review and meta-analysis of prospective studies. J Diabetes Complications 2016;30:368–73.

32. Chen CH, Lin CL, Hsu CY, et al. Association Between Type I and II Diabetes With Gallbladder Stone Disease. Front Endocrinol (Lausanne) 2018;9:720.

33. McCormick N, Yokose C, Lu N, et al. Sodium-Glucose Cotransporter-2 Inhibitors vs Sulfonylureas for Gout Prevention Among Patients With Type 2 Diabetes Receiving Metformin. JAMA Intern Med 2024;184:650–660.

34. Pradhan R, Lu S, Yin H, et al. Novel antihyperglycaemic drugs and prevention of chronic obstructive pulmonary disease exacerbations among patients with type 2 diabetes: population based cohort study. BMJ 2022;379:e071380.

35. Haeusler RA, Pratt-Hyatt M, Welch CL, et al. Impaired generation of 12-hydroxylated bile acids links hepatic insulin signaling with dyslipidemia. Cell Metab 2012;15:65–74.

36. Portincasa P, Di Ciaula A, Wang HH, et al. Coordinate regulation of gallbladder motor function in the gut-liver axis. Hepatology 2008;47:2112–26.

