## supplementary materials for "SGLT2 inhibitors improved clinical outcomes on biliary diseases beyond glycemic control in patients with type 2 diabetes"

**Supplementary Tables**

**Table S1.** Diagnosis codes for all diseases in the study.

**Table S2.** Age-adjusted Charlson Comorbidity Index.

**Table S3.** Individual drug for study medication.

**Table S4.** Sequences of the real-time PCR primers used in this study.

**Table S5.** Key resources table.

**Table S6.** Patient characteristics before and after IPTW with 1% removal.

**Table S7.** Risk of biliary disease for SGLT2i compared with SU group at different time stages.

**Table S8.**Risk of biliary disease for SGLT2i compared with SU group at different time in subgroups analyses.

**Table S9.** Differences in laboratory examinations between the two treatment groups in the index date and the termination date of follow-up.

**Table S10.** Grade of experimental gallstones.

**Table S11.** Serum, bile, and liver biochemistry in LD-fed mice and ND-fed mice.

**Table S12.** List of bile acids.

**Table S13.** KEGG pathways of bile acid metabolomics analysis.

**Supplementary Figures**

**Figure S1.** The distribution of variables after IPTW.

**Figure S2.** The weight, food intake, water intake, liver index, incidence of gallstones, blood glucose, islet function in mice.

**Figure S3.** Ileum feces metabolomics analysis.

**Figure S4.** Colon feces metabolomics analysis.

**Table S1. Diagnosis codes for all diseases in the study**

|  | **ICD-9/10-CM** |
| --- | --- |
| Type 2 diabetes mellitus | E11 |
| Diabetes retinopathy | E113 |
| Diabetes neuropathy | E114 |
| Diabetes nephropathy | E112 |
| Stroke | E13.500x241+I79.2* |
| Metabolic emergency | E110, E111 |
| Cardiovascular disease | E11.502+I79.2* |
| Obstructive sleep apnea-hypopnea syndrome | G47.300x037, G47.301 |
| Hypertension | I10-I16 |
| Hyperlipidemia | E78 |
| Thyroid disease | E01-E07 |
| Vascular disease | I51.600, I67.800, I73.800, I73.900, I78.800, I78.900 |
| Diabetes with organ damage | E112-E117 |
| Chronic kidney disease | N18.800 |
| Congestive heart failure | I50 |
| Ischemic heart disease | E11.500x031+I43.8*, I25.500 |
| Chronic obstructive pulmonary  disease | J44 |
| Any tumor | C00-D48 |
| Cerebral vascular disease | I67.800, |
| Metastatic solid tumor | M80000/6 |
| Leukemia | C91-C95 |
| Hemiplegia | G81 |
| Lymphoma | C81-C88 |
| Dementia | F03 |
| Mild liver disease | K72.905 |
| Connective tissue disease | M35 |
| Ulcer disease | K25, K26, K28 |
| Moderate or severe renal disease | N19.x01, |
| Moderate or severe liver disease | K72.1, K70.300, K74.100, |
| Acquired immune deficiency syndrome | B22, B23, B24 |
| Biliary stones | K80, K80.0, K80.1, K80.2, K80.3, K80.4, K80.5, K80.8 |
| Cholecystitis | K81, K81.0, K81.1, K81.8, K81.9 |
| Other diseases of the gallbladder | K82, K82.0, K82.1, K82.2, K82.3, K82.4, K82.8, K82.9 |
| Other diseases of biliary tract | K83, K83.0, K83.1, K83.2, K83.3, K83.4, K83.5, K83.8, K83.9 |
| Biliary acute pancreatitis | K85.0 |
| Diseases of gallbladder and biliary tract caused by diseases classified elsewhere | K87*, K87.1* |
| Biliary cirrhosis | K74.3, K74.4, K74.5 |
| Cholangiocarcinoma | C22.1, C24.0 |

Abbreviation: ICD-9/10-CM, International Classification of Diseases, Ninth or Tenth Revision, Clinical Modification.

**Table S2. Age-adjusted Charlson Comorbidity Index**

|  | **Conditions** |
| --- | --- |
| Assigned weights for disease |  |
| 1 | Myocardial infarction (MI)  Congestive heart failure (CHF)  Peripheral vascular disease (PVD)  Cerebrovascular disease  Dementia  Chronic pulmonary disease (COPD)  Connective tissue disease (CTD)  Ulcer disease  Mild liver disease (MLD)  Diabetes |
| 2 | Hemiplegia  Moderate or severe renal disease  Diabetes with endo organ damage  Any tumor  Leukemia  Lymphoma |
| 3 | Moderate or severe liver disease (SLD) |
| 6 | Metastatic solid tumor  Acquired immune deficiency syndrome (AIDs) |
| Assigned weights for age |  |
| 1 | For each decade over age 40 years (up to 4 points) |

**Table S3. Individual drugs for study medication**

| **Drug class** | **Generic name of drug** |
| --- | --- |
| Sodium-dependent glucose transporters 2 inhibitors | Canagliflozin, Empagliflozin, Dapagliflozin |
| Sulfonylureas | Glipizide, Gliclazide, Gliquidone, Glimepiride, Glyburide |
| Non-sulfonylureas | Repaglinide, Mitiglinide, Nateglinide |
| Biguanides | Metformin |
| Thiazolidinediones | Troglitazone, Rosiglitazone, Pioglitazone |
| Alpha-glycosidase inhibitors | Acarbose, Miglitol, Voglibose |
| Dipeptidyl peptidase-4 inhibitors | Sitagliptin, Vildagliptin, Saxagliptin, Alogliptin, Linagliptin |
| Glucagon-like peptide-1 receptor agonists | Semaglutide, Dulaglutide, Exenatide, Liraglutide, Loxenatide |
| Insulins | Rapid insulins, Short insulins, Intermediate insulin, Long-acting insulins or insulin analogue |

**Table S4. Sequences of the real-time PCR primers used in this study.**

|  | **Forward** | **Reverse** | **Source** |
| --- | --- | --- | --- |
| Mouse |  |  |  |
| *β-actin* | GTGACGTTGACATCCGTAAAGA | GCCGGACTCATCGTACTCC | Tsingke Biotech |
| *Abcg5* | AATGCTGTGAATCTGTTTCCCA | CCACTTATGATACAGGCCATCCT | Tsingke Biotech |
| *Abcg8* | TGCCCACCTTCCACATGTC | ATGAAGCCGGCAGTAAGGTAGA | Tsingke Biotech |
| *Cyp7a1* | AGCAACTAAACAACCTGCCAGTACTA | GTCCGGATATTCAAGGATGCA | Tsingke Biotech |
| *Cyp7b1* | CAATCCATGCAGTCACCTTC | TGCCTAGAGAAAAACAGAAAGACA | Tsingke Biotech |
| *Cyp8b1* | TACACTCAGCCAGCACCAAG | AAAGAGGCTGTCCTCATGCC | Tsingke Biotech |
| *Cyp27a1* | GCCTCACCTATGGGATCTTCA | TCAAAGCCTGACGCAGATG | Tsingke Biotech |
| *Ldlr* | GGATGGCTATACCTACCCCTCAA | CACATCGTCCTCCAGGCTG | Tsingke Biotech |
| *Asbt* | GTGGGCTTCCTCTGTCAGTT | GCATCATTCCAAGGGCAAGC | Tsingke Biotech |
| *Ostα* | CTGAAGGACACCCCGATGAG | CCTGGGTCATAGATGCCGTC | Tsingke Biotech |
| *Ostβ* | ATCGAAAGAAGCAGCCACAAG | ATGGGGTACTCTCAACGCTC | Tsingke Biotech |
| *Cck* | CGCAGCCGGTAGTCCCTGCAGAA | CCATCCAGCCCATGTAGTCCCGG | Tsingke Biotech |
| *Gastrin* | TCCCTCTCTCCTTTCCTC | CTTCTTCCTCCATTCGTG | Tsingke Biotech |
| **Human** |  |  |  |
| *β-ACTIN* | TGGCACCCAGCACAATGAA | CTAAGTCATAGTCCGCCTAGAAGCA | Tsingke Biotech |
| *LDLR* | TGACTCAGACGAACAAGGCTG | ATCTAGGCAATCTCGGTCTCC | Tsingke Biotech |
| *ABCG5* | AGAGTCAGGATGGCCTGTAT | ATGCTGAGCAGGGCCACTAT | Tsingke Biotech |
| *ABCG8* | GCACTGGTCATGGCTGAGAA | CACAGGAGTCTTGGCTGCTA | Tsingke Biotech |
| *CYP7A1* | AGGACTTCACTCTACACC | GCAGTCGTTACATCATCC | Tsingke Biotech |
| *CYP7B1* | TTCCTCCACTCATACACAATG | CGTGCTTTTCTTCTTACCATC | Tsingke Biotech |
| *CYP27A1* | AGGGCAAGTACCCAATAAGAGA | TCGTTTAAGGCATCCGTGTAGA | Tsingke Biotech |
| *CYP8B1* | ATCGCCTGAAGCCCGTGCAG | AGCTGGGGAGAGGAAGGAGTGC | Tsingke Biotech |

**Table S5.** Key resources table.

|  | **Source** |
| --- | --- |
| C57BL/6J mice | HANGZHOU ZIYUAN LABORATORY ANIMAL TECHNOLOGY CO.,LTD. (CHINA) |
| Lithogenic feed | JIANGSU XIETONG PHARMACEUTICAL BIO-ENGINEERING CO.，LTD. (CHINA) |
| henagliflozin | JIANGSU XIETONG PHARMACEUTICAL BIO-ENGINEERING CO.，LTD. (CHINA) |
| glimepiride | SANOFI BEIJING PHARMACEUTICALS CO., LTD. (CHINA) |
| Ultrasonic diagnostic instruments | Philips iU Elite and Philips EPIQ7, Ultrasound system, Philips Medical System, Bothell, WA, USA. |
| sulfated Cholecystokinin Octapeptide | MCE |
| fetal bovine serum | Gibco, New York, USA |
| T-1095 | SUN-SHINE CHEMICAL TECHNOLOGY CO., LTD. (CHINA) |
| RNA-easy Isolation Reagent | Vazyme, Cat#R701 |
| Cham Q SYBR qPCR Master Mix | Vazyme, Cat#Q341 |
| Graphpad prism | GraphPad, La Jolla, CA, USA |
| R software | Version x64 4.1.3 |

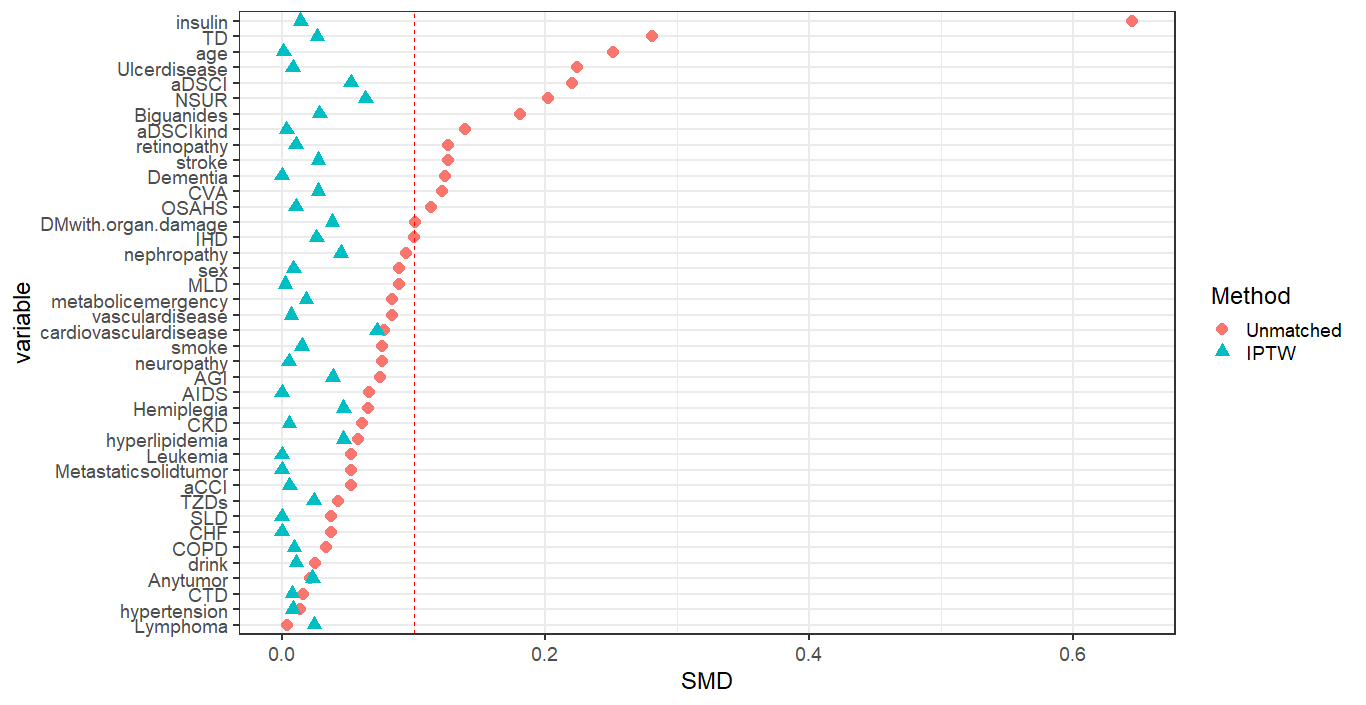

**Figure S1. The distribution of variables after IPTW**

Abbreviations: TD, thyroid disease; aDCSI, the adapted diabetes complications and severity index; NSUR, non-sulfonylureas; CVA, cerebral vascular disease; OSAHS, obstructive sleep apnea-hypopnea syndrome; IHD, ischemic heart disease; MLD, mild liver disease; AGI, alpha-glycosidase inhibitors; AIDS, acquired immune deficiency syndrome. CKD, chronic kidney disease; aCCI, the age-adjusted charlson comorbidity index; TZDs: thiazolidinediones; SLD, moderate or severe liver disease; CHF, congestive heart failure; COPD, chronic pulmonary disease; CTD, connective tissue disease; IPTW, inverse probability of treatment weighting; SMD, standardized mean differences. SMD>0.1 indicates a non-negligible difference between the two treatment groups.

| **Table S6. Patient characteristics before and after IPTW with 1% removal** | | | | | | | | |
| --- | --- | --- | --- | --- | --- | --- | --- | --- |
|  | **before IPTW with 1% removal** | | | | **after IPTW with 1% removal** | | | |
|  | **SGLT2i** | **SU** | **p** | **ASMD** | **SGLT2i** | **SU** | **p** | **ASMD** |
| n | 453 | 1448 |  |  | 441 | 1420 |  |  |
| Sex, Male, n (%) | 307(67.8) | 920(63.5) | 0.112 | 0.089 | 297 (67.3) | 905 (63.7) | 0.892 | 0.009 |
| Age, mean (SD) | 59.48(10.3) | 61.95(9.4) | <0.001 | 0.251^a^ | 59.64(10.3) | 62.04(9.2) | 0.983 | 0.001 |
| Smoke, n (%) | 195(43.0) | 569(39.3) | 0.172 | 0.076 | 191(43.3) | 560(39.4) | 0.812 | 0.015 |
| Drink, n (%) | 143(31.6) | 474(32.7) | 0.685 | 0.025 | 139(31.5) | 464(32.7) | 0.867 | 0.011 |
| Diabetic co-medication, n (%) | | | | | | | | |
| Insulin  Biguanides  TZDs  AGI  NSUR | 336(74.2)  353(77.9)  36(7.9)  199(43.9)  51(11.3) | 637(44.0)  1014(70.0)  99(6.8)  690(47.7)  82(5.7) | <0.001  0.001  0.485  0.183  <0.001 | 0.645^a^  0.181^a^  0.042  0.075  0.202^a^ | 325(73.7)  344(78.0)  36(8.2)  196(44.4)  46(10.4) | 622(43.8)  994(70.0)  98(6.9)  678(47.7)  79(5.6) | 0.828  0.680  0.698  0.549  0.279 | 0.014  0.029  0.024  0.039  0.063 |
| Diabetic complications, n (%) | | | | | | | | |
| aDCSI group | 284(62.7) | 809(55.9) | 0.012 | 0.139^a^ | 273(61.9) | 787(55.4) | 0.956 | 0.004 |
| Diabetic retinopathy | 50(11.0) | 107(7.4) | 0.018 | 0.126^a^ | 45(10.2) | 104(7.3) | 0.863 | 0.010 |
| Diabetic neuropathy | 43(9.5) | 107(7.4) | 0.177 | 0.076 | 39(8.8) | 101(7.1) | 0.931 | 0.005 |
| Diabetic nephropathy | 43(9.5) | 100(6.9) | 0.086 | 0.094 | 35(7.9) | 93(6.5) | 0.403 | 0.045 |
| Stroke | 228(50.3) | 638(44.1) | 0.022 | 0.126^a^ | 220(49.9) | 619(43.6) | 0.668 | 0.028 |
| Metabolic emergency | 14(3.1) | 26(1.8) | 0.137 | 0.084 | 12(2.7) | 25(1.8) | 0.745 | 0.018 |
| Cardiovascular disease | 1(0.2) | 11(0.8) | 0.355 | 0.077 | 1(0.2) | 10(0.7) | 0.156 | 0.072 |
| OSAHS | 9(2.0) | 10(0.7) | 0.032 | 0.113^a^ | 7(1.6) | 9(0.6) | 0.835 | 0.010 |
| Other comorbidities, n (%) | | | | | | | | |
| aCCI, mean (SD) | 5.71(2.1) | 5.60(1.9) | 0.318 | 0.053 | 5.67(2.0) | 5.55(1.8) | 0.926 | 0.006 |
| Hypertension  Hyperlipidemia | 344(75.9)  181(40.0) | 1091(75.3)  620(42.8) | 0.847  0.307 | 0.014  0.058 | 332(75.3)  176(39.9) | 1065(75.0)  605(42.6) | 0.897  0.469 | 0.008  0.047 |
| TD | 171(37.7) | 360(24.9) | <0.001 | 0.281^a^ | 162(36.7) | 345(24.3) | 0.655 | 0.027 |
| Vascular disease | 16(3.5) | 31(2.1) | 0.136 | 0.084 | 15(3.4) | 29(2.0) | 0.889 | 0.007 |
| DM with organ damage | 111(24.5) | 294(20.3) | 0.066 | 0.101^a^ | 103(23.4) | 283(19.9) | 0.527 | 0.038 |
| CKD | 5(1.1) | 8(0.6) | 0.360 | 0.061 | 4(0.9) | 7(0.5) | 0.908 | 0.005 |
| CHF | 0(0.0) | 1(0.1) | 1 | 0.037 | 0(0.0) | 0(0.0) | NA | <0.001 |
| IHD | 222(49.0) | 637(44.0) | 0.069 | 0.101^a^ | 215(48.8) | 619(43.6) | 0.680 | 0.026 |
| COPD | 9(2.0) | 36(2.5) | 0.665 | 0.034 | 9(2.0) | 32(2.3) | 0.895 | 0.009 |
| Any tumor | 28(6.2) | 97(6.7) | 0.780 | 0.021 | 28(6.3) | 89(6.3) | 0.737 | 0.023 |
| CVA | 228(50.3) | 641(44.3) | 0.027 | 0.122^a^ | 220(49.9) | 619(43.6) | 0.668 | 0.028 |
| Metastatic solid tumor | 0(0.0) | 2(0.1) | 1 | 0.053 | 0(0.0) | 0(0.0) | NA | <0.001 |
| Leukemia | 0(0.0) | 2(0.1) | 1 | 0.053 | 0(0.0) | 0(0.0) | NA | <0.001 |
| Hemiplegia | 9(2.0) | 17(1.2) | 0.285 | 0.065 | 432(98.0) | 9(2.0) | 15(1.1) | 0.047 |
| Lymphoma | 2(0.4) | 6(0.4) | 1 | 0.004 | 2(0.5) | 5(0.4) | 0.563 | 0.025 |
| Dementia | 0(0.0) | 11(0.8) | 0.132 | 0.124^a^ | 0(0.0) | 0(0.0) | NA | <0.001 |
| MLD | 20(4.4) | 40(2.8) | 0.109 | 0.089 | 18(4.1) | 37(2.6) | 0.964 | 0.002 |
| CTD | 2(0.4) | 8(0.6) | 1 | 0.016 | 2(0.5) | 8(0.6) | 0.892 | 0.008 |
| Ulcer disease | 166(36.6) | 381(26.3) | <0.001 | 0.224^a^ | 157(35.6) | 371(26.1) | 0.891 | 0.008 |
| SLD | 0(0.0) | 1(0.1) | 1 | 0.037 | 0(0.0) | 0(0.0) | NA | <0.001 |
| AIDS | 1(0.2) | 0(0.0) | 0.539 | 0.067 | 0(0.0) | 0(0.0) | NA | <0.001 |

Abbreviation: IPTW, inverse probability of treatment weighting; ASMD, absolute standardized mean difference; SGLT2i, sodium-glucose cotransporter-2 inhibitors; SU, sulfonylureas; n, number; TZDs, thiazolidinediones; AGI, alpha-glycosidase inhibitors; NSUR, non-sulfonylureas; aDCSI, the adapted diabetes complications and severity index; aCCI, the age-adjusted charlson comorbidity index; OSAHS, obstructive sleep apnea-hypopnea syndrome; SD, standard deviation; TD, thyroid disease; DM, diabetes mellitus; CKD, chronic kidney disease; CHF, congestive heart failure; IHD, ischemic heart disease; COPD, chronic pulmonary disease; CVA, cerebral vascular disease; MLD, mild liver disease; CTD, connective tissue disease; SLD, moderate or severe liver disease; AIDS, acquired immune deficiency syndrome. ^a^ASMD > 0.1 which indicates a non-negligible difference between the two treatment groups.

**Table S7. Risk of biliary disease for SGLT2i compared with SU group at different time stages**

|  | **Patients** | **Disease** | **Incidence rate**  **(per 1000 person-year)** | **HR (95%CI)** | **p** |
| --- | --- | --- | --- | --- | --- |
| **Outcomes at 6 months**  **SGLT2i**  **SU**  **Outcomes at 12 months**  **SGLT2i**  **SU**  **Outcomes at 18 months**  **SGLT2i**  **SU**  **Outcomes at 24 months**  **SGLT2i**  **SU** | 441  1420  441  1420  441  1420  441  1420 | 14  72  16  73  20  134  21  160 | 15.14  17.05  17.30  16.82  21.62  30.88  22.70  36.88 | 0.912(0.475-1.632)  Reference  1.028(0.559-1.784)  Reference  0.7(0.414-1.125)  Reference  0.616(0.371-0.974)  Reference | 0.751  0.919  0.129  0.032^*^ |
| **p for trend** |  |  |  |  | 0.028^*^ |

Abbreviation: SGLT2i, sodium-glucose cotransporter-2 inhibitors; SU, sulfonylureas; HR, hazard ratios; CI, confidence intervals. *The asterisk indicates a statistically significant difference.

| **Table S8. Risk of biliary disease for SGLT2i compared with SU group at different time in subgroups analyses** | | | | | | | | | | |
| --- | --- | --- | --- | --- | --- | --- | --- | --- | --- | --- |
|  | **Subgroups** | | | | | | | | | |
| **Follow-up period** | **N** | **Events** | **SGLT2i#** | **SU#** | **HR95%CI (RF:SU)** | **N** | **Events** | **SGLT2i#** | **SU#** | **HR95%CI (RF:SU)** |
|  | Male | | | | | Female | | | | |
| All | 1202 | 155 | 31.15 | 48.40 | 0.644(0.376-1.044) | 659 | 53 | 6.35 | 33.33 | 0.190(0.022-0.724) |
|  | age≤60years | | | | | ＞60years | | | | |
| All | 788 | 84 | 23.86 | 41.72 | 0.572(0.274-1.085) | 1073 | 124 | 21.55 | 44.01 | 0.490(0.229-0.934) |
|  | aDCSI=0(without diabetic complications) | | | | | aDCSI>0(with at least one diabetic complication) | | | | |
| All | 801 | 94 | 19.07 | 44.13 | 0.432(0.169-0.929) | 1060 | 114 | 25.09 | 42.22 | 0.594(0.314-1.044) |
|  | aCCI <5(low group) | | | | | aCCI >/=5 (high group) | | | | |
| All | 562 | 63 | 31.25 | 42.26 | 0.739(0.335-1.469) | 1299 | 145 | 18.18 | 43.42 | 0.419(0.204-0.774) |
|  | Treated with canagliflozin | | | | | Treated with dapagliflozin or empagliflozin | | | | |
| All | 55 | 7 | 7.57 | 43.09 | 0.176(0.070-0.369) | 386 | 14 | 15.14 | 43.09 | 0.351(0.188-0.604) |

\# incidence rate per 1000 person-years; *The asterisk indicates a statistically significant difference;

Abbreviation: SGLT2i, sodium-glucose cotransporter-2 inhibitors; SU, sulfonylureas; HR, hazard ratios; CI, confidence intervals; aDCSI, the adapted diabetes complications and severity index; aCCI, the age-adjusted charlson comorbidity index; RF, reference group.

**Table S9. Differences in laboratory examinations between the two treatment groups in the index date and the termination date of follow-up.**

|  | **SGLT2i** | **p1** | **SU** | **p2** | **t/Z** | **p** |
| --- | --- | --- | --- | --- | --- | --- |
| Weight, M(IQR), kg  Index date  Termination date | 69(60,80)  69(61.75,78) | 0.529 | 69(64,76)  70(64,76) | 0.062 | -0.319  -0.787 | 0.750  0.431 |
| BMI, mean (SD), kg/m^2^  Index date  Termination date | 25.30(3.53)  25.28(2.69) | 0.605 | 25.36(2.86)  25.50(2.37) | 0.056 | -0.505  -1.273 | 0.614  0.203 |
| FBG, M(IQR), mmol/L  Index date  Termination date | 7.81(6.31,10.34)  6.80(5.83,8.50) | 0.000 | 7.64(5.86,10.50)  6.94(5.52,8.71) | 0.000 | -0.577  -0.107 | 0.564  0.915 |
| HbA1c, mean (SD), %  Index date  Termination date | 8.18(1.78)  7.44(1.57) | 0.000 | 7.89(2.10)  7.61(2.00) | 0.000 | 2.279  -1.506 | 0.023^*^  0.132 |
| TC, M(IQR), mmol/L  Index date  Termination date | 4.41(3.70,5.16)  4.32(3.53,5.27) | 0.021 | 4.41(3.76,5.19)  4.29(3.59,5.09) | 0.000 | -0.076  -0.733 | 0.939  0.464 |
| TG, M(IQR), mmol/L  Index date  Termination date | 1.58(1.09,2.46)  1.43(1.05,1.93) | 0.000 | 1.52(1.06,2.21)  1.44(1.02,2.03) | 0.000 | -1.659  -0.17 | 0.097  0.865 |
| LDLC, mean (SD), mmol/L  Index date  Termination date | 2.65(0.89)  2.58(0.94) | 0.093 | 2.67(0.93)  2.55(0.94) | 0.000 | -0.425  0.483 | 0.671  0.629 |
| HDLC, mean (SD), mmol/L  Index date  Termination date | 1.08(0.20)  1.13(0.22) | 0.000 | 1.11(0.29)  1.08(0.30) | 0.000 | -1.852  3.121 | 0.064  0.002^***^ |
| LPa, M(IQR), mg/L  Index date  Termination date | 42(15.82,150)  37(12,94) | 0.002 | 91(29.75,203)  89.2(28.52,194) | 0.512 | -4.308  -6.715 | 0.000^***^  0.000^***^ |
| TBA, M(IQR), μmol/L  Index date  Termination date | 3.40(2.40,6.02)  2.60(2.10,4.13) | 0.000 | 3.80(2.40,6.20)  3.70(2.40,5.85) | 0.011 | -0.693  -5.064 | 0.488  0.000^***^ |
| TBIL, M(IQR), μmol/L  Index date  Termination date | 12.59(9.30,16.40)  11.64(8.80,15.60) | 0.000 | 12.70(9.68,16.50)  12.49(9.30,16.60) | 0.052 | -0.836  -2.459 | 0.403  0.014^*^ |
| DBIL, M(IQR), μmol/L  Index date  Termination date | 4.22(3.40,5.60)  4.10(3.10,5.40) | 0.001 | 4.33(3.20,5.70)  4.20(3.12,5.70) | 0.007 | -0.365  -1.503 | 0.715  0.133 |
| IBIL, M(IQR), μmol/L  Index date  Termination date | 7.40(5.17,10.30)  6.85(4.98,9.60) | 0.002 | 7.30(5.10,9.99)  7.20(5.10,9.80) | 0.436 | -0.530  -0.953 | 0.596  0.341 |

Abbreviation: FPG, fasting plasma glucose; HbA1c, Glycosylated Hemoglobin; SGLT2i, sodium-glucose cotransporter-2 inhibitors; SU, sulfonylureas; TC, total cholesterol; TG, triglyceride; LDLC, low density lipoprotein cholesterol; HDLC, high density lipoprotein cholesterol; LPa, lipoprotein a; TBA, total bile acid; TBIL, total bilirubin; DBIL, direct bilirubin; IBIL, indirect bilirubin; SD, standard deviation; M(IQR), median (interquartile range); N, number.

The analysis was performed on 1861 patients after 1% removal with IPTW. P value is the comparison of clinical characteristics in SGLT2i group and SU group both in index date and termination date. The p1 value compares the change in clinical features from the index date to the termination date in the SGLT2i group, while p2 is in the SU group. *The asterisk indicates a statistically significant difference.

**Table S10. Grade of experimental gallstones**

|  | **Grade of experimental gallstones** | | | | | | **p** |
| --- | --- | --- | --- | --- | --- | --- | --- |
|  | **0** | **I** | **II** | **III** | **IV** | **V** |  |
| ND+placebo | 10 |  |  |  |  |  | <0.001^***^ |
| LD+placebo | 1 | 2 | 2 | 1 | 4 | 1 |  |
| LD+Glim | 2 | 2 | 2 | 2 | 2 | 2 | 0.994 |
| LD+Hena | 4 | 2 | 1 | 2 |  |  | 0.028^*#^ |

*The asterisk and p value indicated a statistically significant difference between different groups and LD+placebo group.

#The pound key indicates a statistically significant difference between LD-fed with SGLT2i and SU treatment.

Abbreviation: ND, normal diet; LD, lithogenic diet; Glim, glimepiride; Hena, henagliflozin.

**Table S11. Serum, bile, and liver biochemistry in LD-fed mice and ND-fed mice.**

|  | **ND+placebo**  **(n=10)** | **LD+placebo**  **(n=12)** | **LD+Glim**  **(n=12)** | **LD+Hena**  **(n=11)** |
| --- | --- | --- | --- | --- |
| FBG, mmol/L | 5.15±0.57 | 5.19±0.74 | 5.91±1.19 | 5.69±1.02 |
| FINS, ng/mL | 0.52±0.29 | 0.35±0.11 | 0.35±0.13 | 0.44±0.35 |
| HOMA-IR | 2.56±1.67 | 1.69±0.51 | 2.04±0.88 | 2.37±1.77 |
| AUC of IPGTT | 1712.40±223.90^*^ | 1434.83±204.34 | 1322.33±215.26 | 1468.45±247.85 |
| liver index | 0.044±0.007^***^ | 0.092±0.006 | 0.087±0.006 | 0.080±0.006^***^ |
| ALT, U/L | 31.90±15.07^***^ | 487.00±268.39 | 320.33±63.04^*^ | 279.27±54.03^**^ |
| AST, U/L | 126.90±29.31^***^ | 330.50±105.65 | 256.50±50.33^*^ | 230.91±44.90^**^ |
| ALP, U/L | 77.40±4.93^***^ | 237.17±32.33 | 220.17±49.05 | 183.64±46.60^**^ |
| GGT, U/L | 1.90±1.66 | 2.58±1.78 | 2.83±1.75 | 3.64±1.63 |
| TBA, umol/L | 36.99±60.74^**^ | 127.65±51.90 | 112.70±83.15 | 55.67±34.32^*^ |
| TBIL, umol/L | 5.92±1.96^**^ | 7.75±0.85 | 7.77±1.03 | 6.51±0.65 |
| DBIL, umol/L | 0.53±0.36 | 0.97±0.72 | 0.88±0.68 | 0.55±0.47 |
| IBIL, umol/L | 5.39±1.90 | 6.78±1.09 | 6.88±1.38 | 5.96±0.62 |
| TC, mmol/L | 2.73±0.62^***^ | 5.61±0.64 | 5.00±0.48 | 4.63±0.64^**^ |
| TG, mmol/L | 0.98±0.14^***^ | 0.57±0.10 | 0.50±0.08 | 0.47±0.08 |
| LDLC, mmol/L | 0.28±0.06^***^ | 1.31±0.13 | 1.28±0.13 | 1.12±0.14^**##^ |
| HDLC, mmol/L | 1.83±0.48 | 1.82±0.19 | 1.63±0.16 | 1.55±0.20 |
| TBA in bile, mmol/L | 944.10±49.87^***^ | 463.52±9.43 | 590.43±14.44^***^ | 720.06±4.08^***###^ |
| TC in bile, mmol/L | 3.17±0.75^*^ | 5.52±1.05 | 5.73±0.56 | 4.85±1.11 |
| LDLC in bile, mmol/L | 1.44±1.01^**^ | 4.60±1.22 | 4.26±0.86 | 2.76±0.43 |
| TC in liver, umol/g | 9.88±1.68^***^ | 44.29±4.90 | 42.80±4.16 | 39.15±2.49^*^ |
| TG in liver, umol/g | 9.70±1.93^***^ | 15.50±3.03 | 12.96±3.07 | 12.34±3.80 |
| LDLC in liver, umol/g | 2.28±1.18^***^ | 13.28±3.41 | 12.13±3.08 | 10.02±1.55^*^ |

Results are expressed as mean±SEM. Statistically significant changes between LD placebo mice and the other three groups are marked ^*^p<0.05, ^**^p<0.01,^***^p<0.001. Statistically significant differences between LD-fed with Hena and Glim treatment are marked ^#^p<0.05, ^##^p<0.01, ^###^p<0.001.

Abbreviation: ND, normal diet; LD, lithogenic diet; Glim, glimepiride; Hena, henagliflozin; FPG, fasting plasma glucose; FINS, fasting insulin; HOMA-IR, homeostasis model assessment of insulin resistance; AUC, area under curve; IPGTT, intraperitoneal glucose tolerance test; ALT, alanine transaminase; AST, aspartate aminotransferase; ALP, alkaline phosphatase; GGT, gamma-glutamyl transpeptidase; TBA, total bile acid; TBIL, total bilirubin; DBIL, direct bilirubin; IBIL, indirect bilirubin; TC, total cholesterol; TG, triglyceride; LDLC, low density lipoprotein cholesterol; HDLC, high density lipoprotein cholesterol; n, number.

**Table S12. List of bile acids**

| **English name** | **Abbreviation** | **CAS** | **formula** |
| --- | --- | --- | --- |
| Dehydrolithocholic acid | DHLCA | 1553-56-6 | C24H38O3 |
| Isoallolithocholic acid/Isolithocholic acid | —/isoLCA | 2276-93-9/1534-35-6 | C24H40O3 |
| Lithocholic acid | LCA | 434-13-9 | C24H40O3 |
| 23-Nordeoxycholic acid | 23norDCA | 53608-86-9 | C23H38O4 |
| 6-Ketolithocholic acid | 6-ketoLCA | 2393-61-5 | C24H38O4 |
| 7-Ketolithocholic acid | 7-ketoLCA | 4651-67-6 | C24H38O4 |
| 12-Ketolithocholic acid | 12-ketoLCA | 5130-29-0 | C24H38O4 |
| Apocholic acid | apoCA | 641-81-6 | C24H38O4 |
| Isoursodeoxycholic acid | isoUDCA | 78919-26-3 | C24H40O4 |
| Murideoxycholic acid | MDCA | 668-49-5 | C24H40O4 |
| Isohyodeoxycholic acid | isoHDCA | 570-84-3 | C24H40O4 |
| Ursodeoxycholic acid | UDCA | 128-13-2 | C24H40O4 |
| Hyodeoxycholic acid | HDCA | 83-49-8 | C24H40O4 |
| 3-Epideoxycholic acid | βDCA | 570-63-8 | C24H40O4 |
| Chenodeoxycholic acid | CDCA | 474-25-9 | C24H40O4 |
| Deoxycholic acid | DCA | 83-44-3 | C24H40O4 |
| Isodeoxycholic acid | isoDCA | 566-17-6 | C24H40O4 |
| Nor Cholic Acid | NCA | 60696-62-0 | C23H38O5 |
| Dehydrocholic acid | DHCA | 81-23-2 | C24H34O5 |
| 7,12-Diketolithocholic acid | 7,12-diketoLCA | 517-33-9 | C24H36O5 |
| 6,7-Diketolithocholic acid | 6,7-diketoLCA | — | C24H36O5 |
| 7-Ketodeoxycholic acid | 7-KHCA | 911-40-0 | C24H38O5 |
| 12-Dehydrocholic acid | 12-DHCA | 2458-8-4 | C24H38O5 |
| 3-Dehydrocholic acid | 3-DHCA | 2304-89-4 | C24H38O5 |
| Ursocholic acid | UCA | 2955-27-3 | C24H40O5 |
| 3β-Cholic Acid | βCA | 3338-16-7 | C24H40O5 |
| ω-Muricholic Acid | ω-MCA | 6830-03-1 | C24H40O5 |
| α-Muricholic acid | α-MCA | 2393-58-0 | C24H40O5 |
| β-Muricholic acid | β-MCA | 2393-59-1 | C24H40O5 |
| Hyocholic acid | HCA | 547-75-1 | C24H40O5 |
| Allocholic acid | ACA | 2464-18-8 | C24H40O5 |
| Cholic acid | CA | 81-25-4 | C24H40O5 |
| Glycolithocholic acid | GLCA | 474-74-8 | C26H43NO4 |
| Glycoursodeoxycholic acid | GUDCA | 64480-66-6 | C26H43NO5 |
| Glycohyodeoxycholic acid | GHDCA | 13042-33-6 | C26H43NO5 |
| Glycochenodeoxycholic acid | GCDCA | 640-79-9 | C26H43NO5 |
| Glycodeoxycholic acid | GDCA | 360-65-6 | C26H43NO5 |
| Lithocholic Acid-3-Sulfate | LCA-3S | 64936-81-8 | C24H40O6S |
| Glycodehydrocholic acid | GDHCA | 3415-45-0 | C26H37NO6 |
| Glycohyocholic acid | GHCA | 32747-08-3 | C26H43NO6 |
| Glycocholic acid | GCA | 475-31-0 | C26H43NO6 |
| Ursodeoxycholic acid 3-Sulfate | UDCA-3S | 68780-73-4 | C24H40O7S |
| Chenodeoxycholic Acid-3-Sulfate | CDCA-3S | 59132-32-0 | C24H40O7S |
| Deoxycholic Acid-3-Sulfate | DCA-3S | 67030-48-2 | C24H40O7S |
| Taurolithocholic acid | TLCA | 6042-32-6 | C26H45NO5S |
| Cholic Acid-3-Sulfate | CA-3S | 58822-34-7 | C24H40O8S |
| Tauroursodeoxycholic acid | TUDCA | 14605-22-2 | C26H45NO6S |
| Taurohyodeoxycholic acid | THDCA | 2958-04-5 | C26H45NO6S |
| Taurochenodeoxycholic acid | TCDCA | 516-35-8 | C26H45NO6S |
| Taurodeoxycholic acid | TDCA | 516-50-7 | C26H45NO6S |
| Glycolithocholic Acid-3-Sulfate | GLCA-3S | 15324-64-8 | C26H43NO7S |
| Tauro ω-muricholic acid | T-ω-MCA | 130325-58-5 | C26H45NO7S |
| Tauro α-Muricholic acid | T-α-MCA | 25613-05-2 | C26H45NO7S |
| Tauro β-Muricholic acid | T-β-MCA | 25696-60-0 | C26H45NO7S |
| Taurohyocholic acid | THCA | 32747-07-2 | C26H45NO7S |
| Taurocholic acid | TCA | 81-24-3 | C26H45NO7S |
| Glycoursodeoxycholic Acid-3-Sulfate | GUDCA-3S | 133429-88-6 | C26H43NO8S |
| Glycochenodeoxycholic Acid 3-Sulfate | GCDCA-3S | 66874-09-7 | C26H43NO8S |
| Glycodeoxycholic Acid-3-Sulfate | GDCA-3S | 66874-10-0 | C26H43NO8S |
| Glycocholic Acid-3-Sulfate | GCA-3S | 67850-84-4 | C26H43NO9S |
| Taurolithocholic Acid-3-Sulfate | TLCA-3S | 15324-65-9 | C26H45NO8S2 |
| Chenodeoxycholic acid-3-β-D-Glucuronide | CDCA-3Gln | 58814-71-4 | C30H48O10 |
| Chenodeoxycholic acid 24-Acyl-β-D-glucuronide | CDCA-24Gln | 208038-27-1 | C30H48O10 |
| Tauroursodeoxycholic Acid-3-Sulfate | TUDCA-3S |  | C26H45NO9S2 |
| Taurochenodeoxycholic Acid-3-Sulfate | TCDCA-3S | 67030-59-5 | C26H45NO9S2 |
| Taurodeoxycholic Acid-3-Sulfate | TDCA-3S |  | C26H45NO9S2 |
| Taurocholic Acid-3-Sulfate | TCA-3S | 67030-62-0 | C26H45NO10S2 |
| Glycodeoxycholic acid-3-O-β-glucuronide | GDCA-3Gln | 75672-36-5 | C32H51NO11 |
| Glycochenodeoxycholic Acid-3-O-β-glucuronide | GCDCA-3Gln | 75672-22-9 | C32H51NO11 |

**Table S13. KEGG pathway of bile acid metabolomics analysis.**

|  | **Description** | **Compounds in serum(all)** | **Compounds in ileum(all)** |
| --- | --- | --- | --- |
| mmu00120 | Primary bile acid biosynthesis - Mus musculus (house mouse) | C00695;C01921;C05465;C05122 | C02528;C00695;C05466;C01921;C05465;C05122 |
| mmu04979 | Cholesterol metabolism - Mus musculus (house mouse) | C01921;C05465;C05122 | C05466;C01921;C05465;C05122 |
| mmu00430 | Taurine and hypotaurine metabolism - Mus musculus (house mouse) | C05122 | C05122 |

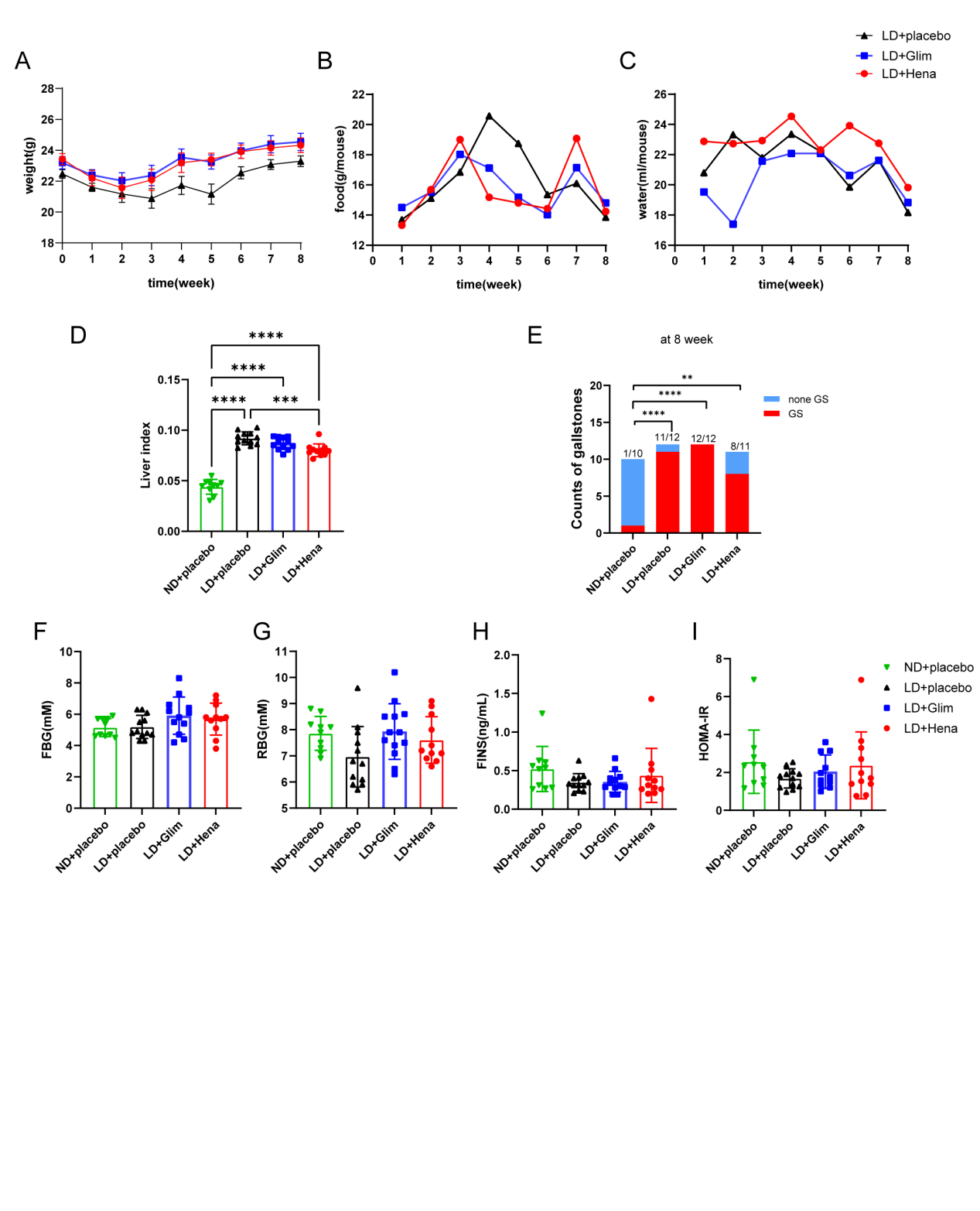

**Figure S2. The weight, food intake, water intake, liver index, incidence of gallstones, blood glucose, and islet function in mice.**

(A) Weight (g) in different groups during 8 weeks of feeding. **(B)** Food intake (g/mouse) in different groups during 8 weeks of feeding. **(C)** Water intake (ml/mouse) in different groups during 8 weeks of feeding. **(D)** Liver index of four groups. **(E)** The incidences of gallstones in four groups after different treatments for 8 weeks. **(F)** FBG (mM), **(G)** RBG (mM), **(H)** Fasting insulin **(**FINS) (ng/mL), **(I)** Homeostasis model assessment (HOMA-IR) in each group, which was calculated by the following formula: FBG*FINS / 22.5.

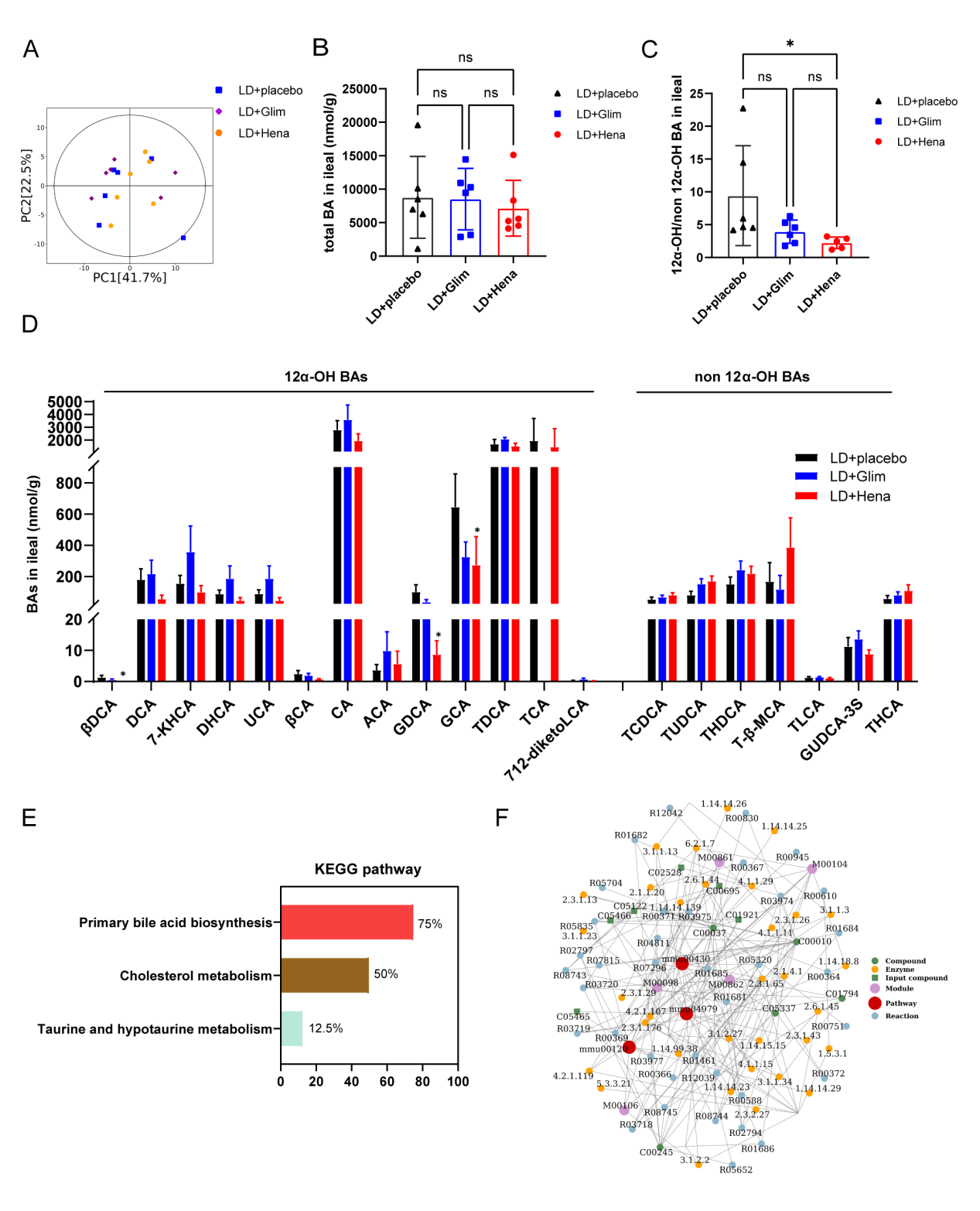

**Figure S3. Ileum feces metabolomics analysis.**

**(A)** Principal component analysis (PCA) in three different LD-fed mice groups after 8 weeks of treatment. **(B)** Total BA in ileum feces. **(C)** 12α-OH/non 12α-OH BAs in ileum feces. **(D)** Relative quantification of specific bile acid components in ileum feces. **(E)** KEGG analysis. **(F)** Network analysis for three groups. The red dots represent a metabolic pathway, and the yellow dots represent relevant regulatory enzyme information.

*The asterisk indicates a statistically significant difference between LD-fed with henagliflozin or glimepiride treatment and LD-fed with placebo mice. *p <0.05.

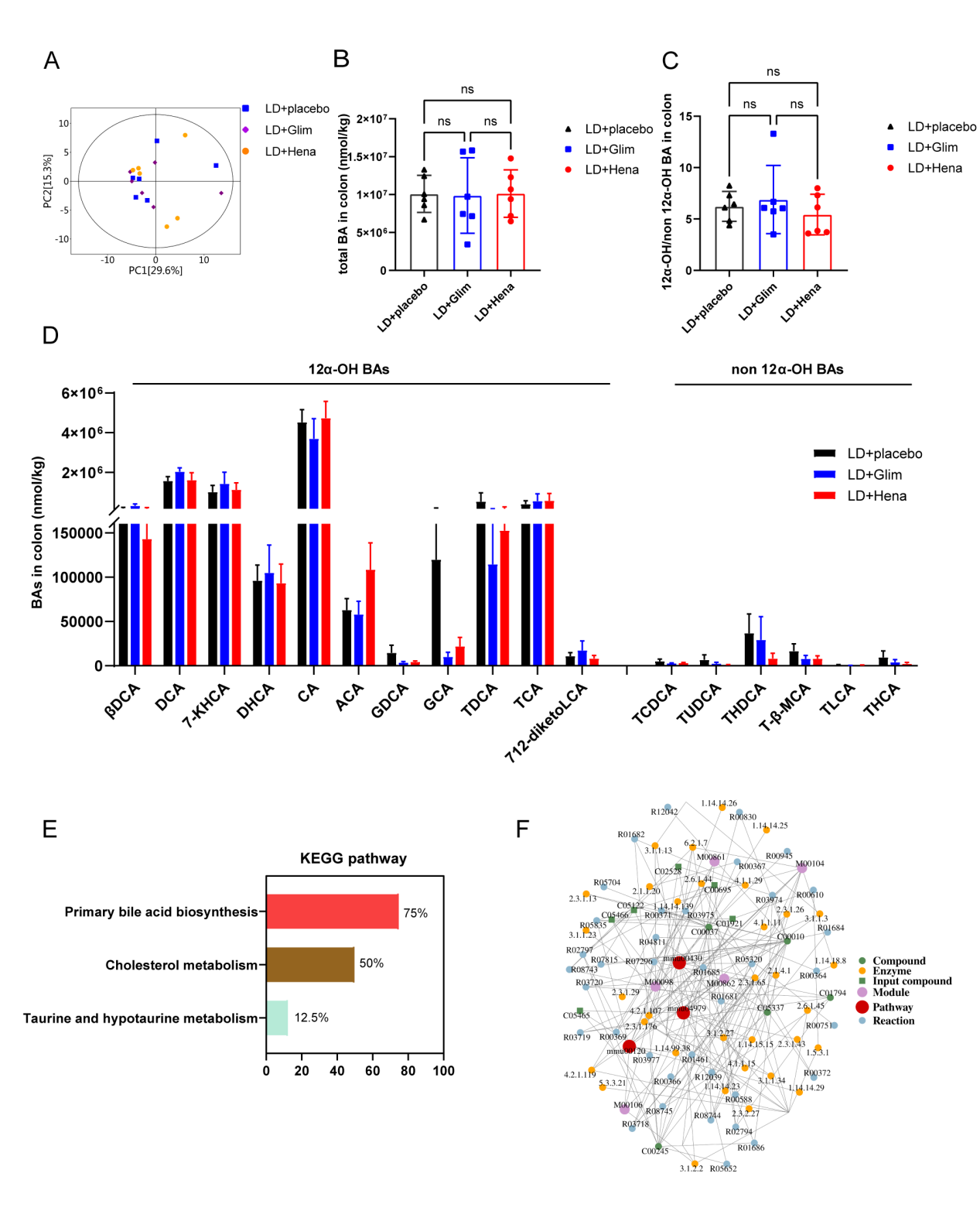

**Figure S4. Colon feces metabolomics analysis.**

**(A)** Principal component analysis (PCA) in three different LD-fed mice groups after 8 weeks of treatment. **(B)** Total BA in colon feces. **(C)** 12α-OH/non 12α-OH BAs in colon feces. **(D)** Relative quantification of specific bile acid components in colon feces. **(E)** KEGG analysis. **(F)** Network analysis for three groups. The red dots represent a metabolic pathway, and the yellow dots represent relevant regulatory enzyme information.

*The asterisk indicates a statistically significant difference between LD-fed with henagliflozin or glimepiride treatment and LD-fed with placebo mice. *p <0.05.
